# Completeness of reporting of clinical prediction models developed using supervised machine learning: A systematic review

**DOI:** 10.1101/2021.06.28.21259089

**Authors:** Constanza L Andaur Navarro, Johanna A A Damen, Toshihiko Takada, Steven W J Nijman, Paula Dhiman, Jie Ma, Gary S Collins, Ram Bajpai, Richard D Riley, Karel GM Moons, Lotty Hooft

## Abstract

**Objective:** While many studies have consistently found incomplete reporting of regression-based prediction model studies, evidence is lacking for machine learning-based prediction model studies. We aim to systematically review the adherence of Machine Learning (ML)-based prediction model studies to the Transparent Reporting of a multivariable prediction model for Individual Prognosis Or Diagnosis (TRIPOD) Statement.

**Study design and setting:** We included articles reporting on development or external validation of a multivariable prediction model (either diagnostic or prognostic) developed using supervised ML for individualized predictions across all medical fields (PROSPERO, CRD42019161764). We searched PubMed from 1 January 2018 to 31 December 2019. Data extraction was performed using the 22-item checklist for reporting of prediction model studies (www.TRIPOD-statement.org). We measured the overall adherence per article and per TRIPOD item.

**Results:** Our search identified 24 814 articles, of which 152 articles were included: 94 (61.8%) prognostic and 58 (38.2%) diagnostic prediction model studies. Overall, articles adhered to a median of 38.7% (IQR 31.0-46.4) of TRIPOD items. No articles fully adhered to complete reporting of the abstract and very few reported the flow of participants (3.9%, 95% CI 1.8 to 8.3), appropriate title (4.6%, 95% CI 2.2 to 9.2), blinding of predictors (4.6%, 95% CI 2.2 to 9.2), model specification (5.2%, 95% CI 2.4 to 10.8), and model’s predictive performance (5.9%, 95% CI 3.1 to 10.9). There was often complete reporting of source of data (98.0%, 95% CI 94.4 to 99.3) and interpretation of the results (94.7%, 95% CI 90.0 to 97.3).

**Conclusion:** Similar to prediction model studies developed using conventional regression-based techniques, the completeness of reporting is poor. Essential information to decide to use the model (i.e. model specification and its performance) is rarely reported. However, some items and sub-items of TRIPOD might be less suitable for ML-based prediction model studies and thus, TRIPOD requires extensions. Overall, there is an urgent need to improve the reporting quality and usability of research to avoid research waste.

**What is new?:**

- **Key findings:** Similar to prediction model studies developed using regression techniques, machine learning (ML)-based prediction model studies adhered poorly to the TRIPOD statement, the current standard reporting guideline.
- **What this adds to what is known?** In addition to efforts to improve the completeness of reporting in ML-based prediction model studies, an extension of TRIPOD for these type of studies is needed.
- **What is the implication, what should change now?** While TRIPOD-AI is under development, we urge authors to follow the recommendations of the TRIPOD statement to improve the completeness of reporting and reduce potential research waste of ML-based prediction model studies.

## INTRODUCTION

Clinical prediction models are used extensively in healthcare to aid patient diagnosis and prognosis of disease and health status. A diagnostic model combines multiple predictors or test results to predict the presence or absence of a certain disorder, whereas a prognostic model estimates the probability of future occurrence of an outcome.^1–3^ Studies developing, validating, and updating prediction models are abundant in most clinical fields and their number will continue to increase as prediction models developed using artificial intelligence (AI) and machine learning (ML) are receiving substantial interest in the healthcare community.^4^

ML, a subset of AI, offers a class of models that can iteratively learn from data, identify complex data patterns, automate model building, and predict outcomes based on what has been learned using computer-based algorithms.^5,6^ ML is often described as more efficient and accurate than conventional regression-based techniques. ML-based prediction models, correctly developed, validated, and implemented, can improve patient benefit, and reduce disease and health system burden. There is increasing concern of the methodological and reporting quality of studies developing prediction models, with research till date focusing on models developed with conventional statistical techniques such as logistic and Cox regression.^7–11^ Recent studies have found limited application of ML-based prediction models because of poor study design and reporting.^12,13^

Incomplete (or unclear) reporting makes ML-based prediction models difficult to interpret and impedes validation by independent researchers, thus creating barriers to their use in daily clinical practice. Complete and accurate reporting of ML-based prediction model studies will improve its interpretability, reproducibility, risk of bias assessment, and applicability in daily medical practice and is, therefore, essential for high-quality research.^14^ To improve transparency and reporting of prediction model studies, the Transparent Reporting of a multivariable prediction model for Individual Prognosis Or Diagnosis (TRIPOD) Statement, a checklist of 22 items, was designed (www.tripod-statement.org).^15,16^ Specific guidance for ML-based prediction model studies is currently lacking and has initiated the extension of TRIPOD for prediction models developed using ML or AI (TRIPOD-AI).^17^

We conducted a systematic review to assess the completeness of reporting of ML-based diagnostic and prognostic prediction model studies in recent literature using the TRIPOD Statement.^15,16^ Our results will highlight specific reporting areas that can inform reporting guidelines for ML, such as TRIPOD-AI. ^17^

## METHODS

Our systematic review protocol was registered (PROSPERO, CRD42019161764) and published.^18^ We reported this systematic review following the PRISMA statement.^19^

### Data source and search

We searched PubMed on December 19, 2019 to identify primary articles describing prediction models (diagnostic or prognostic) using any supervised ML technique across all clinical domains published between 1 January 2018 and 31 December 2019. The search strategy is provided in the supplemental material.

### Study selection

We included articles that described the development or validation of one or more multivariable prediction models using any supervised ML technique aiming for individualized prediction of risk or outcomes. As there is still no consensus on a definition of ML, we defined a ‘study using ML’ as a study that describes the use of a non-generalized linear models to develop or validate a prediction model (e.g. tree-based models, ensembles, deep learning). Hence, studies that claimed to have used ML, but they reported only regression-based statistical techniques were excluded from this systematic review (e.g. logistic regression, lasso regression, ridge regression and elastic net). Specifically, we focused on supervised ML, a subdomain of ML, that is characterized by using an algorithm that learns to predict from labelled outcome examples. Example are random forest, support vector machine, neural network, naïve bayes, and gradient boosting.

Articles reporting on the incremental value or model extension were also included. We included all articles regardless of study design, data source, or patient-related health outcome. Articles that investigated a single predictor, test or biomarker, or its causality with an outcome were excluded. Articles using ML to enhance reading of images or signals, or articles where ML models only used genetic traits or molecular markers as predictors, were also excluded. We also excluded systematic reviews, conference abstracts, tutorials, and articles for which full-text was unavailable via our institution. We restricted the search to human subjects and English-language articles. Further details are stated in our protocol.^18^

Two researchers, from a group of seven (CLAN, TT, SWJN, PD, JM, RB, JAAD), independently screened titles and abstracts to identify potentially eligible studies. Full-text articles were then retrieved, and two independent researchers reviewed them for eligibility using Rayyan.^20^ One researcher (CLAN) screened all articles and six researchers (TT, SWJN, PD, JM, RB, JAAD) collectively screened the same articles. Disagreements between reviewers were resolved by a third researcher (JAAD).

### Data extraction

The data extraction form was based on the TRIPOD adherence assessment form (www.tripod-statement.org).^21^ This form contains several adherence statements (hereafter called sub-items) per TRIPOD item. Some items and sub-items are applicable to all types of studies, while others are only applicable to model development only or external validation only (Table 1). To judge reporting of the requested information, sub-items were formulated to be answered with ‘yes’, ‘no’, ‘not applicable’. We amended the published adherence form by omitting the ‘referenced’ option because we checked the information in the references, supplemental material or appendix. Sub-items related to items 10b and 16 were extracted per model, rather than at study-level, as they refer to model performance.

**Table 1.** TRIPOD adherence reporting items

| Reporting Items |  | Study design | If applicable to studies | Reporting items for TRIPOD adherence |  |
| --- | --- | --- | --- | --- | --- |
|  |  |  |  | Development only | Development and validation |
| **1. Title** |  | D,V |  | ✓ | ✓ |
| **2. Abstract** |  | D,V |  | ✓ | ✓ |
| **Introduction** |  |  |  |  |  |
| **3. Background and objectives** | a. Context and rationale | D,V |  | ✓ | ✓ |
|  | b. Objectives | D,V |  | ✓ | ✓ |
| **Methods** |  |  |  |  |  |
| **4. Source of data** | a. Source of data | D,V |  | ✓ | ✓ |
|  | b. Key dates | D,V |  | ✓ | ✓ |
| **5. Participants** | a. Study setting | D,V |  | ✓ | ✓ |
|  | b. Eligibility criteria | D,V |  | ✓ | ✓ |
|  | c. Details of treatment | D,V | ✓ | ✓ | ✓ |
| **6. Outcome** | a. Outcome definition | D,V |  | ✓ | ✓ |
|  | b. Blinding of outcome assessment | D,V |  | ✓ | ✓ |
| **7. Predictors** | a. Predictors definition | D,V |  | ✓ | ✓ |
|  | b. Blinding of predictor assessment | D,V |  | ✓ | ✓ |
| **8. Sample size** | Arrival at study size | D,V |  | ✓ | ✓ |
| **9. Missing Data** | Handling of missing data | D,V |  | ✓ | ✓ |
| **10. Statistical analysis** | a. Handling of predictors in the analysis | D |  | ✓ | ✓ |
|  | b. Specification of the model, all model building procedures, and internal validation methods | D |  | ✓ | ✓ |
|  | c. For validation, description of how predictions were calculated | V |  | ✓ | n.a. |
|  | d. Specification of all measures used to assess model performance | D,V |  | ✓ | ✓ |
|  | e. Description of model updating | V | ✓ | ✓ | n.a. |
| **11. Risk groups** | Details of how risk groups were created | D,V | ✓ | ✓ | ✓ |
| **12. Development vs. validation** | For validation, description of differences between development and validation data | V |  | ✓ | ✓ |
| **Results** |  |  |  |  |  |
| **13. Participants** | a. Flow of participants through the study | D,V |  | ✓ | ✓ |
|  | b. Description of characteristics of participants | D,V |  | ✓ | ✓ |
|  | c. For validation, comparison with development data | V |  | ✓ | ✓ |
| **14. Model development** | a. Number of participants and outcome in each analysis | D |  | ✓ | ✓ |
|  | b. Unadjusted association between each candidate predictor and outcome | D | ✓ | ✓ | ✓ |
| **15. Model specification** | a. Presentation of full prediction model | D | ✓ | ✓ | ✓ |
|  | b. Explanation of how to use the prediction model | D |  | ✓ | ✓ |
| **16. Model performance** | Report of model performance measures | D,V |  | ✓ | ✓ |
| **17. Model updating** | Results from any model updating | V | ✓ | ✓ | n.a. |
| **Discussion** |  |  |  |  |  |
| **18. Limitations** | Limitations | D,V |  | ✓ | ✓ |
| **19. Interpretation** | **a.** For validation, interpretation of performance measure results | V |  |  | ✓ |
|  | **b.** Overall interpretation of results | D,V |  | ✓ | ✓ |
| **20. Implications** | Potential clinical use of the model and implications for future research | D,V |  | ✓ | ✓ |
| **Other information** |  |  |  | ✓ | ✓ |
| **21. Supplementary information** | Availability of supplementary resources | D,V |  | ✓ | ✓ |
| **22. Funding** | Source of funding and role of funders | D,V |  | ✓ | ✓ |
| **Total number of applicable items for TRIPOD adherence score** |  |  |  | **31** | **37** |

(n.a) No included studies reported external validation only or model updating (Item 10c, 10e, and 17)

We performed a double data extraction for included articles. Two reviewers independently extracted data from each article using the standardized form which was available in REDCap, a data capture tool.^22^ To accomplish consistent data extraction, the form was piloted by all reviewers on five articles. One researcher (CLAN) extracted data from all articles and six researchers (TT, SWJN, PD, JM, RB, JAAD) collectively extracted data from the same articles. Discrepancies in data extraction were discussed and resolved between each pair of reviewers.

### Data synthesis and analysis

We categorized prediction model studies as prognosis or diagnosis and classified studies by research aim: development (with or without internal validation), development with external validation (same model), development with external validation (different model), and external validation only. Detailed definition of research aims can be found in the supplemental material. Where articles described the development and/or validation of more than one prediction model, we chose the first ML model reported in the methods section for extraction.

We scored each TRIPOD item as ‘reported’ and ‘not reported’ based on answers to corresponding sub-items. If the answer to all sub-items of a TRIPOD item is scored ‘yes’ or ‘not applicable’, the corresponding item was considered ‘reported’. Two analyses were conducted: adherence per item and overall adherence per article. We calculated the adherence per TRIPOD item by dividing the number of studies that adhered to a specific item by the number of studies in which the item was applicable. The total number of TRIPOD items varies by the type of prediction model study (Table 1). We calculated the overall adherence to TRIPOD per article by dividing the sum of reported TRIPOD items by the total number of applicable TRIPOD items for each study. If an item was ‘not applicable’ for a particular study, it was excluded when calculating the overall adherence, both in the numerator and denominator.^21^ Analyses were performed using R version 3.6.2 (R Core Team, Vienna, Austria). Results were summarized as percentages, medians, ranges, and using visual plots.

## RESULTS

We identified 24 814 unique articles, of which we sampled ten random sets of 249 articles each with sampling replacement for screening. We screened the title and abstracts of 2 482 articles, screened full-text of 312 articles and included 152 eligible articles (Figure 1).

**Figure 1.**
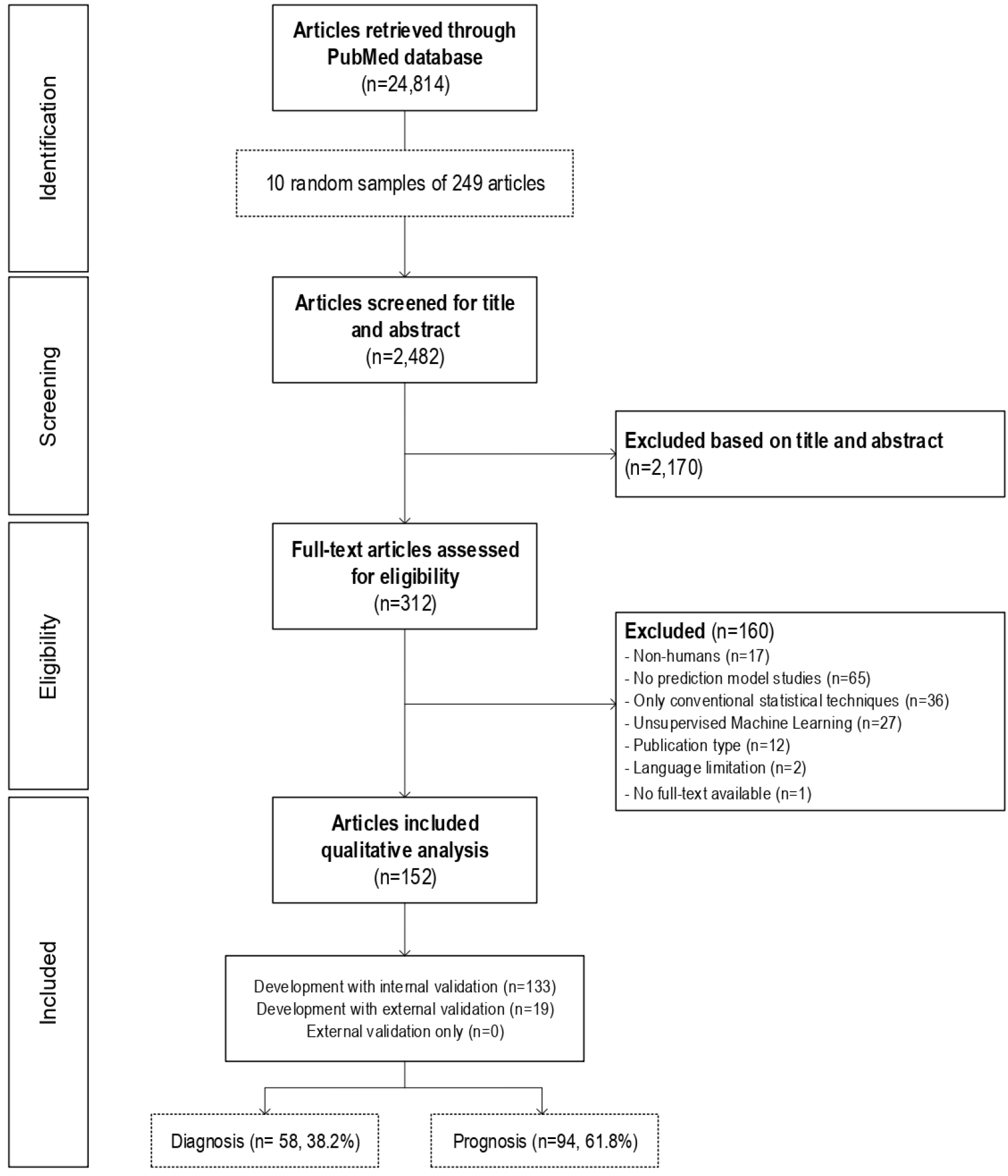
Flowchart of included studies

We included 94 (61.8%) prognostic and 58 (38.2%) diagnostic prediction model studies. 132 (86.8%) articles described development with internal validation and 19 (12.5%) development with external validation (same model). One (0.6%) article was development with external validation (different model) and was included as a development with internal validation study in the present analysis. Prediction models were developed most often in oncology (21/152 [13.8%]). Detailed description of the included studies is provided in supplemental material.

Across the 152 studies, 1429 models were developed and 219 were validated, with a range of 1 to 156 for both types of studies. The most commonly used ML techniques for the first reported model were Classification and Regression Tree (CART [10.1%]), Support Vector Machine (SVM [9.4%]) and Random Forest (RF [9.4%]). Alongside ML techniques, 19.5% of studies reported the development of a model using conventional statistical techniques, such as logistic regression. Five out of 152 studies (3.3%, 95% CI 1.4% to 7.5%) stated following the recommendations of the TRIPOD Statement.

### Overall adherence per TRIPOD item

Five TRIPOD items reached at least 75% adherence (background, objectives, source of data, limitations, and interpretation), whilst 12 TRIPOD items were below 25% adherence (Figure 2). Results for the overall adherence per TRIPOD item stratified by study type, diagnosis and prognosis, and publication year are shown in Table 2.

**Figure 2.**
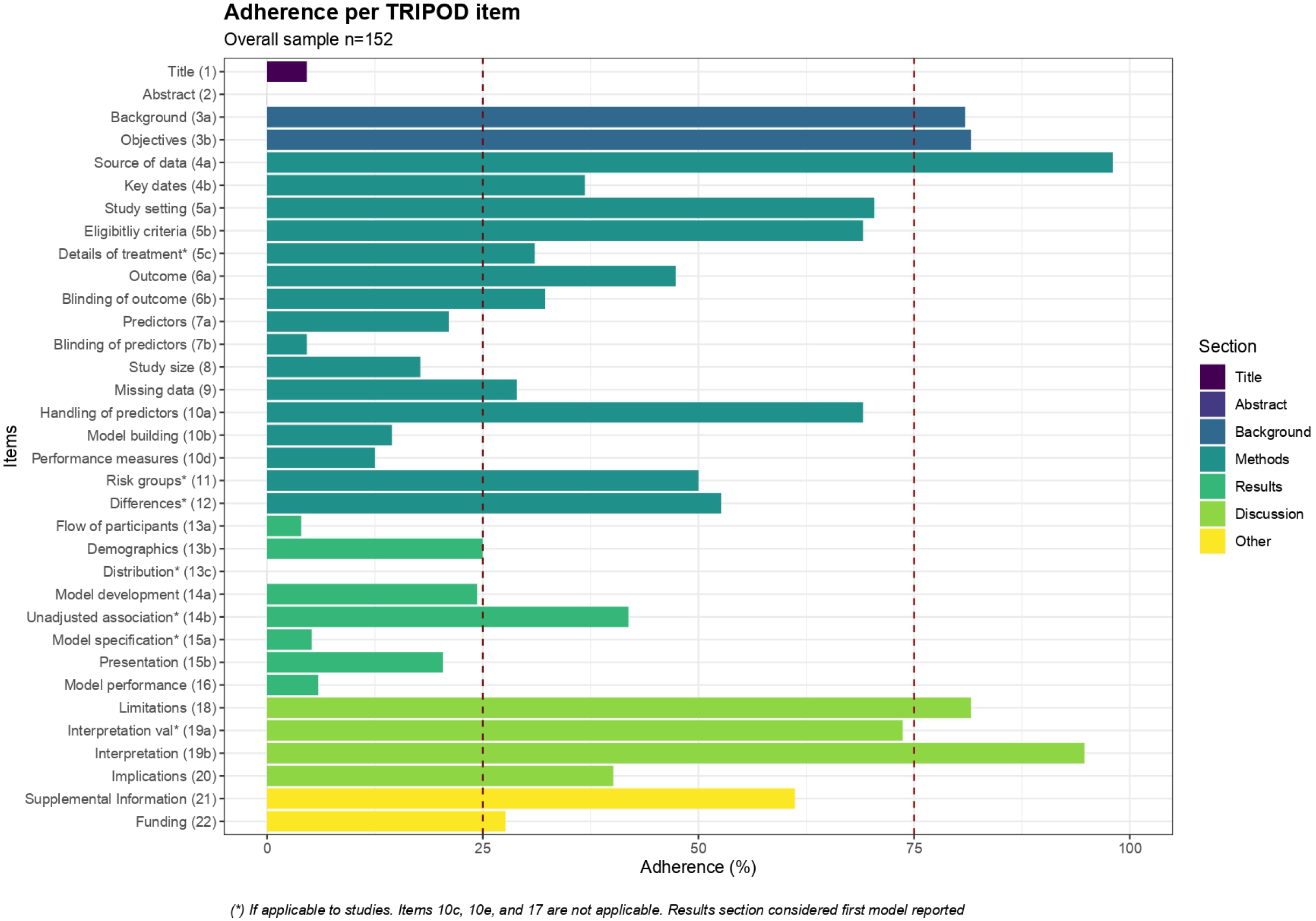
Overall adherence per TRIPOD item

**Table 2.**
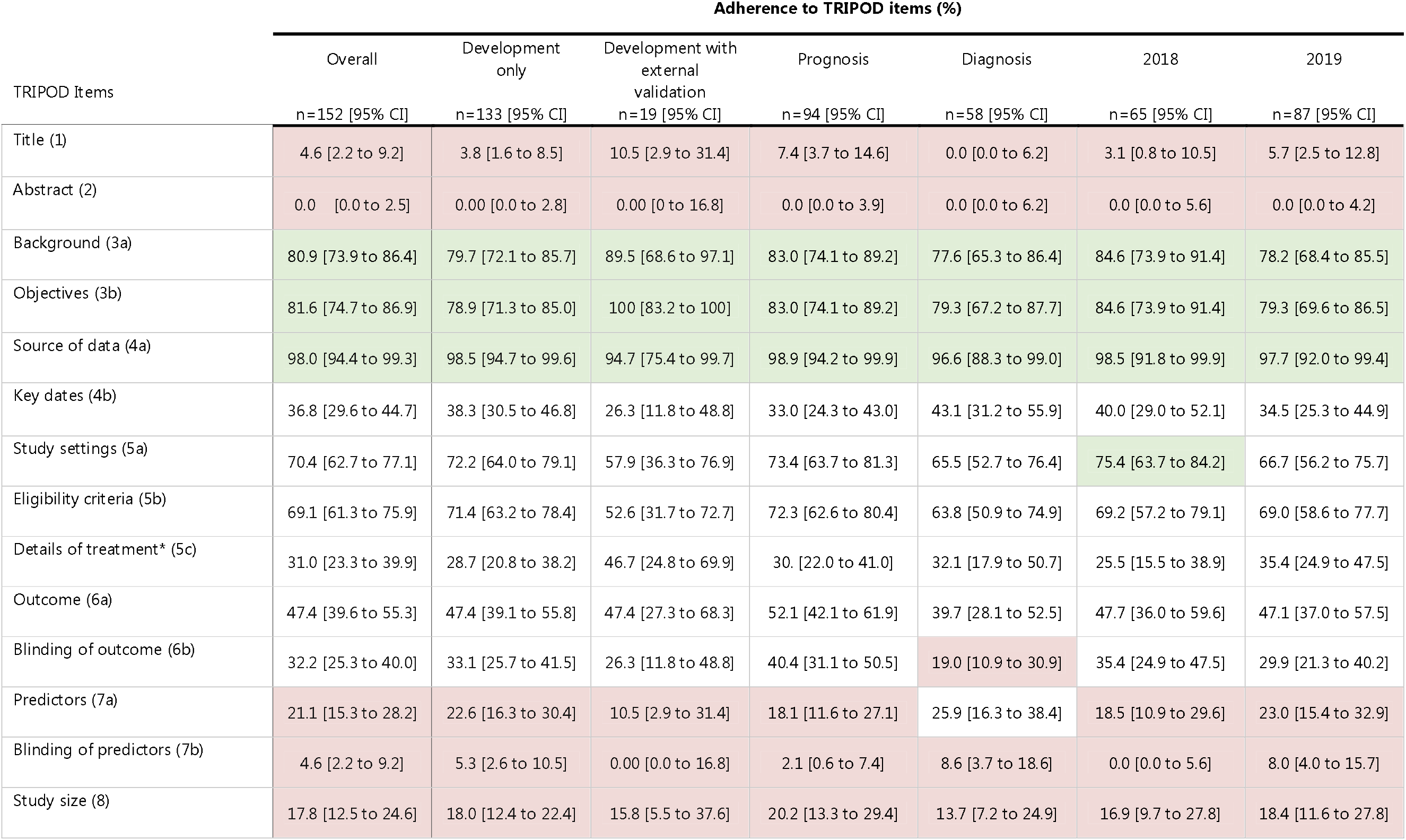
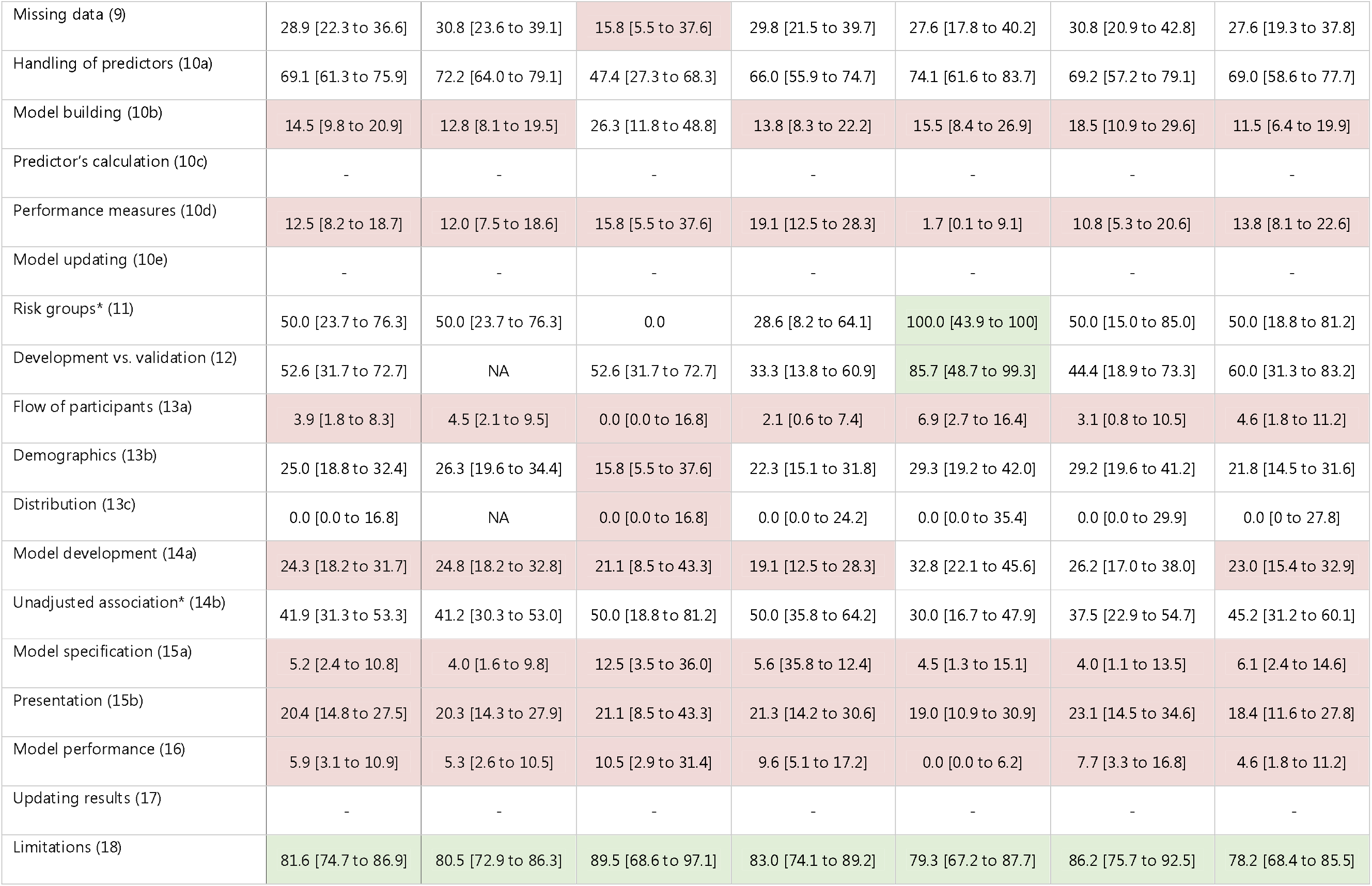
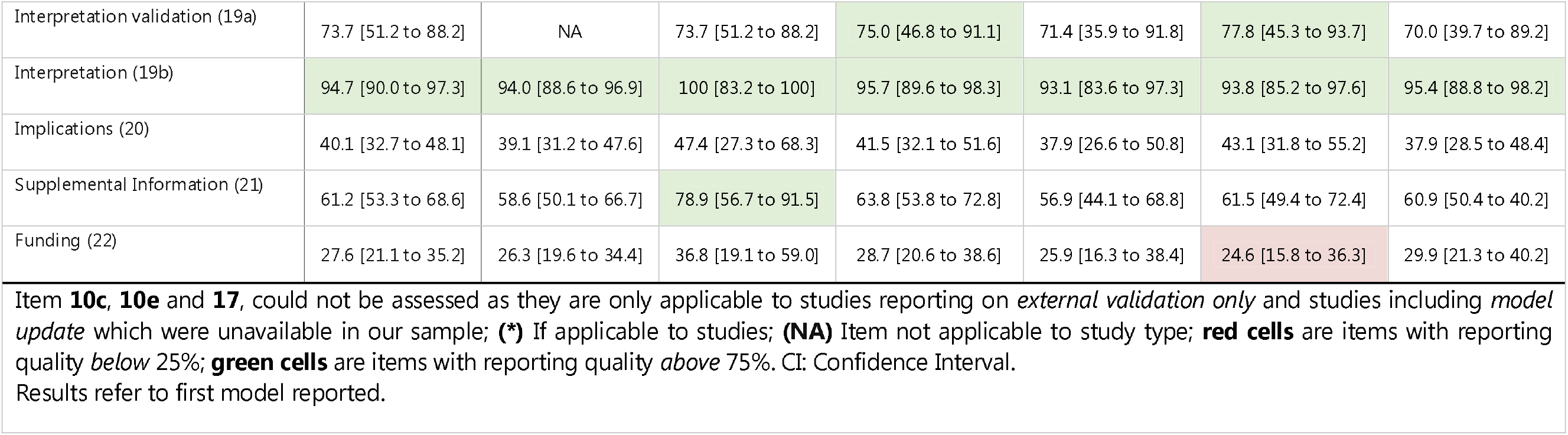
Adherence to TRIPOD items

#### Title and abstract (item 1 and 2)

Seven out of 152 studies (4.6%, 95% CI 2.2 to 9.2) completely adhered to title recommendations. Description of type of prediction model study (sub-item 1.i) was poorly reported (11.2%, CI 7.0 to 17.2), but outcome to be predicted (sub-item 1.iv) was well reported (91.4%, CI 85.9 to 94.9). No study fully reported item 2, abstract (0.0%, CI 0.0%to 2.5).

#### Introduction (item 3)

Background and objectives were most often reported TRIPOD items. Background was provided in 123 studies (80.9%, 95% CI 73.9 to 86.4), and the objectives were reported in 124 studies (81.6%, CI 74.6 to 86.9).

#### Methods (item 4-12)

Source of data was the most often reported item in the methods section, and across all TRIPOD items (98.0%, 95% CI 94.4 to 99.3). Study setting was reported in 107 studies (70.4%, CI 62.7 to 77.1), eligibility criteria in 105 (69.1%, CI 61.3 to 75.9), and handling of predictors in 105 out of 152 studies (69.1%, CI 61.3 to 75.9). Ten studies assessed risk groups and five reported complete information (50.0%, CI 23.7 to 76.3). Differences between development and validation set were reported in 10 out of 19 applicable studies (52.6%, CI 31.7 to 72.7). For 72 studies, definition of outcome was reported (47.4%, CI 39.6% to 55.3). Key study dates such as start and end date of accrual, and length of follow-up were completely reported in 56 studies (36.8%, CI 29.6 to 44.7). Details of treatment were reported in 36 out of applicable 116 studies (31.0%, CI 23.3 to 39.9). Blinding of outcome and predictors were reported in 49 (32.2%, CI 25.3 to 40.0) and 7 studies (4.6%, CI 2.2 to 9.2), respectively.

Forty-four studies reported how missing data were handled (28.9%, 95% CI 22.3 to 36.6). The missing data item consists of four sub-items of which three were rarely addressed in included studies. Within 28 studies that reported handling of missing data: three studies reported the software used (10.7%, CI 3.7 to 27.2), four studies reported the variables included in the procedure (14.3%, CI 5.7 to 31.5) and no study reported the number of imputations (0.0%, CI 0.0 to 39.0). Predictor definitions were given in 32 out of 152 studies (21.1%, CI 15.3 to 28.2), and justification of study size was reported in 27 studies (17.8%, CI 12.5 to 24.6). Model building procedures, such as predictor selection and internal validation, were reported in 22 out of 152 studies (14.5%, CI 9.8 to 20.9). Internal validation, a sub-item of item 10b, was one of the most reported sub-items across studies (91.4%, CI 85.9 to 94.9).

Reporting of measures used to assess and quantify the predictive performance was complete in 19 studies (12.5%, 95% CI 8.2 to 18.7). Though 106 studies (69.7%, CI 62.0 to 76.5) reported discrimination (sub-item 10d.i), only 19 studies (12.5%, CI 8.2 to 18.7) reported calibration (sub-item 10d.ii). Definitions of discrimination and calibration are stated in supplemental material. Other performance measures (sub-item 10d.iii), for example sensitivity, specificity, or predictive values, were reported in 124 studies (81.6%, CI 74.7 to 86.9).

### Results (item 13-17)

Study participant characteristics were reported in 38 out of 152 studies (25.0%, 95% CI 18.8 to 32.4). Basic demographics, at least age and gender (sub-item 13b.i), were provided in 117 studies (77.0%, CI 69.7 to 83.0), while summary information of the predictors (sub-item 13b.ii) was reported in 67 studies (44.1%, CI 36.4 to 52.0). Number of study participants with missing data for predictors (sub-item 13b.iii) was reported in 15 studies (24.2%, CI 15.2 to 36.2). Unadjusted associations were reported in 41 out of the 74 studies that reported regression-based models alongside with ML-models (41.9%, CI 31.3 to 53.3). The number of participants and events were described in 37 studies (24.3%, CI 18.2 to 31.7). In 31 out of 152 studies, an explanation on how to use the developed model to make predictions for new individuals was provided, often in the form of a scoring rule or online calculator (20.4%, CI 14.8 to 27.5). Flow of participants was reported in 6 studies (3.9%, CI 1.8 to 8.3) and model specification was reported in 6 out of 116 applicable studies (5.2%, CI 2.4 to 10.8). Model predictive performance was completely reported in 9 out of 152 studies (5.9%, CI 3.1 to 10.9).

#### Discussion (items 18-20)

Overall interpretation of results was reported in 124/152 studies (81.6%, 95% CI 74.7 to 86.9). Limitations of the study were reported in 144 studies (94.7%, CI 90.0 to 97.3). An interpretation of model performance in the validation set in comparison with the development set was given in 14/19 studies (73.7%, CI 51.2 to 88.2). Potential clinical use and implications for future research was reported in 61 studies (40.1%, CI 32.7 to 48.1).

#### Other information (items 21 and 22)

Availability of supplementary resources was mentioned in 93/152 studies (61.2, 95% CI 53.3 to 68.6). Funding information was reported in 42 studies (27.6%, CI 21.1 to 35.2).

### Overall adherence per article

Overall adherence of studies to items of the TRIPOD Statement ranged between 13.0% and 65.0%; median adherence was 38.7% (IQR 31.0 to 46.5). The completeness reporting in prognostic model studies was higher (median adherence=40.0% (IQR 33.3 to 46.8)) than diagnostic model studies (median adherence=35.7% (IQR 30.2 to 45.0)) (Figure 3). Moreover, median adherence was 40.6% (CI 28.6 to 46.1) in development (with internal validation) studies, compared to 37.9% (CI 31.0 to 46.4) in development with external validation studies.

**Figure 3.**
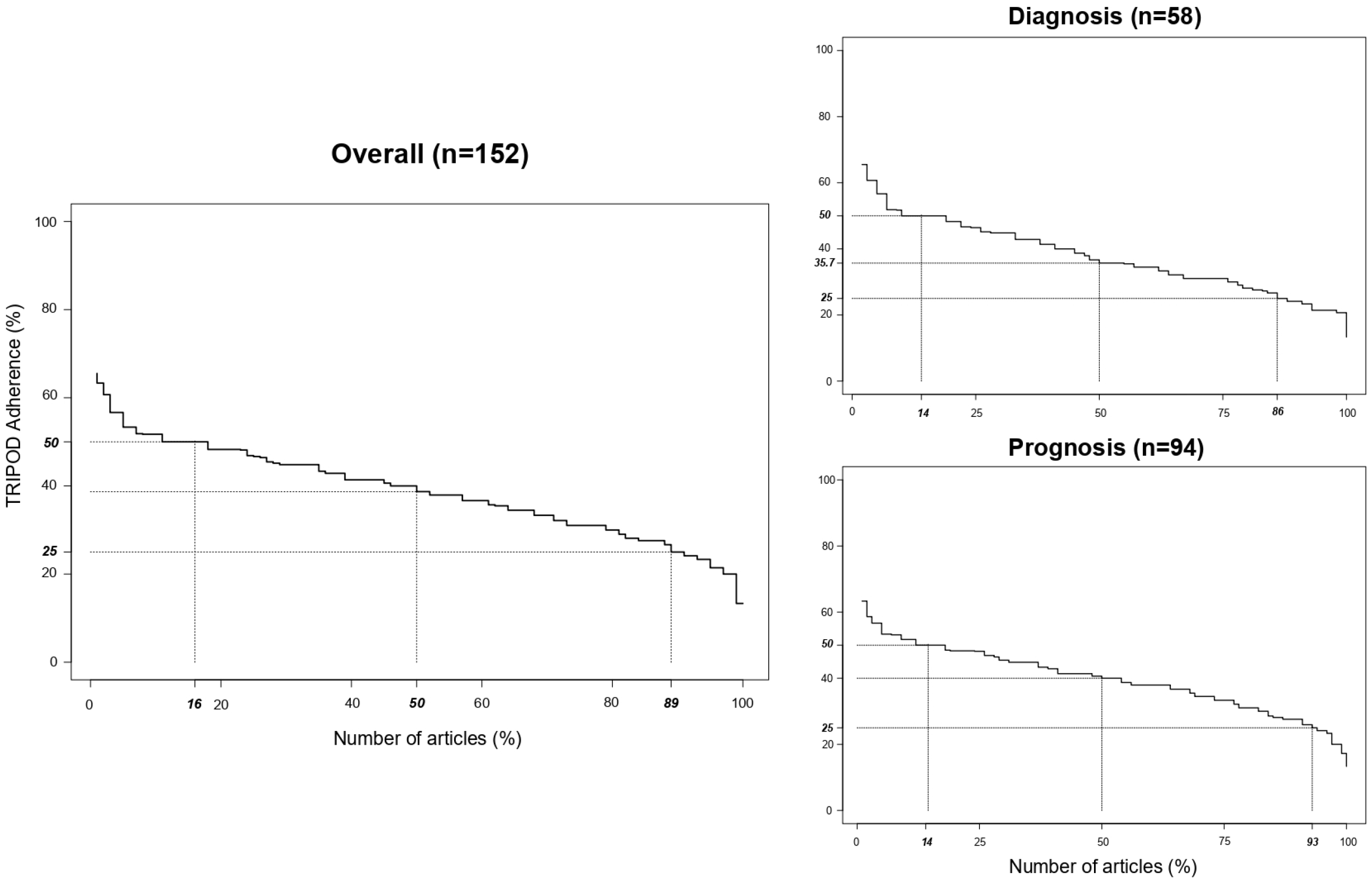
Overall adherence per article

## DISCUSSION

We conducted a systematic review of ML-based diagnostic and prognostic prediction model studies and assessed their adherence to the TRIPOD Statement. We found that ML-based prediction model studies adhere poorly to the TRIPOD Statement reporting items.

Complete reporting in titles and abstracts is crucial to identify and screen articles. However, titles and abstracts were fully reported in less than 5% of articles. In addition, information about methods was infrequently reported. Complete and accurate reporting of the methods used to develop or validate a prediction model facilitates external validation, as well as replication of study results by independent researchers. For example, to enhance transparency and risk of bias assessment, it is recommended to report the number of participants with missing data and report how missing data were handled in the analysis. Handling of missing data was seldom reported, but this may be partially explained by the fact that some ML techniques can handle missing data by design (e.g. sparsity aware splitting in XGBoost and surrogate splits in decision trees).^23,24^ Also most studies divided a single dataset into three: training, validation and test set; the last is used for internal validation. The split sample approach for internal validation was among the most reported sub-items in our sample, but several methodological studies and guidelines have long discouraged this approach.^25^ We included diagnostic model studies that used images as one of the predictors, and deep learning. Often, these studies use several numerical variables based on pixels or voxels and build prediction models based on several layers of statistical interaction. Both topics are challenging to report due to number of variables used and poor interpretability of interactions. This may explain why diagnostic ML-based model studies were slightly worse reported compared to prognostic studies.

Overall, most articles adhered to less than half of the applicable items considered essential for complete reporting. Authors may have avoided reporting specific details about methods and results because their objective may be to explore the data and modeling technique accuracy, rather than build models for individualized predictions in “real world” clinical settings. However, high-quality reporting is also essential for reproducibility and replication. Also, most developed models were unavailable for replication, assessment, or clinical application. Only five studies reported using the TRIPOD Statement for reporting their research. Although TRIPOD was published and disseminated in 2015, it is infrequently used for reporting of ML-based prediction model studies.

Previous systematic reviews have shown poor reporting of regression-based prediction model studies. ^7,8,10^ One study assessed the completeness of reporting of articles published in high impact journals during 2014 within 37 different clinical fields. In 146 studies, over half of TRIPOD items were not fully reported, obtaining an overall adherence of 44% (IQR 35.0 to 52.0). Comparable to our study, the review found poor reporting of the title, abstract, model building, model specification and model performance.^7^ A recent study assessed the completeness of reporting of deep learning-based diagnostic model studies. Although they developed their own data extraction for reporting quality, authors found poor reporting of demographics, distribution of disease severity, patient flow, and distribution of alternative diagnosis.^26^ These items were also inappropriately reported in our study with a median adherence between 0.0% and 47.3%. Another systematic review that assessed studies comparing the performance of diagnostic deep learning algorithms for medical imaging versus expert clinicians reported the overall adherence to TRIPOD was poor with a median of 62.0% (45.0 to 69.0).^27^ In line with our results, a study about the performance of ML models showed that 68.0% of included articles had unclear reporting.^12^

To our knowledge, this is the first systematic review evaluating the completeness of reporting of supervised ML-based prediction model studies in a broad sample of articles. We ran a validated search strategy and performed paired screening. We also used a contemporary sample of studies in our review (2018-2019). Though some eligible articles may have been missed, it is unlikely they would change the conclusions of this review.

We used a systematic scoring-system enhancing the objectivity and consistency for the evaluation of adherence to a reporting guideline.^21^ We used the formal TRIPOD adherence form and checklist for data extraction and assessment; however, these were developed for studies developing prediction models with regression techniques. Although we applied the option ‘not applicable’ for items that were unrelated to ML and items were excluded when calculating overall adherence, our results should be interpreted within this context.

While some items and sub-items may be less relevant for prediction models developed with ML techniques, other items are more relevant for transparent reporting in these studies. For example, source of data (4a), study size (8), missing data (9), transformation of predictors (10a.i), internal validation (10b.iv), and availability of the model (15b) acquire new relevance within the context of ML-based prediction model studies. As ML techniques are prone to overfitting, we recommend to extend item 10b of the TRIPOD adherence form to include a new sub-item specifically related to penalization or shrinkage techniques. New reporting items such as the hardware (i.e. technical aspects) that was used to develop or validate an algorithm in images studies are needed, as well as data clustering. New practices such as explaining models through feature importance plot or tuning of hyper-parameters could be also added to the extension of TRIPOD for ML-based prediction models. Items such as testing of interaction terms (Item 10b-iv), unadjusted associations (14b), and regression coefficients (15a) require updating. Despite these recommendations, most TRIPOD items and sub-items are still applicable for both, regression and ML techniques and should be used to improve reporting quality.

We identified nearly 25 000 articles with prediction and ML-related terms within 2 years, similar to previous systematic reviews about deep learning models.^28,29^ The literature has become saturated with ML-based studies; thus, their identification, reporting and assessment becomes even more relevant. If studies are presented without essential details to make predictions in new patients, subsequent researchers will develop a new model, rather than validating or updating an existing model. Reporting guidelines aim to increase the transparent evaluation, replication and translation of prediction models into clinical practice.^30^ Some reporting guidelines for ML clinical prediction models have been developed. ^31,32^ However, these guidelines are limited and do not follow the EQUATOR recommendations for developing consensus-based reporting guidelines.^33^ The improvement in reporting after the introduction of a guideline has shown to be slow.^30^ Improving the completeness of reporting of ML-based studies might be even more challenging given the number of techniques and associated details that need to be reported. There are also practical issues, like terminology used, word limits, or journal requirements, that are acting as barriers to complete reporting. To overcome these barriers, the use of online repositories for data, script, and complete pipeline could help researchers share their models with enough details to make predictions in new patients and to allow external validation of the model. Our results will provide input and support for the development of TRIPOD-AI, an initiative launched in 2019.^17^ We call for a collaborative effort between algorithm developers, researchers, and journal editors to improve the adoption of good scientific practices related to reporting quality.

## CONCLUSION

ML-based prediction model studies currently do not adhere well to the TRIPOD reporting guideline. More than half of the TRIPOD items considered essential for transparent reporting were inadequately reported, especially regarding details of title, abstract, blinding, model building procedures, model specifications and model performance. Whilst ML brings new challenges to the development of tailored reporting guidelines, our study serves as a baseline measure to define future updates or extensions of TRIPOD tailored to ML modelling strategies.

## Supporting information

PRISMA Statement

## Data Availability

Detailed extracted data on all included studies are available upon reasonable request.

## Contributors

The study concept and design were conceived by CLAN, JAAD, PD, LH, RDR, GSC, and KGMM. CLAN, JAAD, TT, SN, PD, JM and RB conducted article screening and data extraction. CLAN performed data analysis and JAAD verified the underlying data. CLAN wrote the first draft of this manuscript, which was critically revised for important intellectual content by all authors who have provided the final approval of this version. CLAN, the corresponding author, is the guarantor of the review. The corresponding author attests that all listed authors meet authorship criteria and that no others meeting the criteria have been omitted.

## Disclosures

GSC, RDR and KGMM are members of the TRIPOD Group. All authors have nothing to disclose.

## Data sharing

The study protocol is available at doi: 10.1136/bmjopen-2020-038832. The search strategy is available in appendix; detailed extracted data are available upon reasonable request.

## Acknowledgements

The authors would like to thank and acknowledge the support of René Spijker, information specialist.

## Funding support

This study did not receive any specific grant from funding agencies in the public, commercial, or not-for-profit sectors. GSC is supported by the National Institute for Health Research (NIHR) Oxford Biomedical Research Centre (BRC) and by Cancer Research UK program grant (C49297/A27294). PD is supported by the NIHR Oxford BRC. The views expressed are those of the authors and not necessarily those of the NHS, NIHR, or Department of Health.

## Ethical approval

Not required.

## Notes

### Competing Interest Statement

The authors have declared no competing interest.

### Funding Statement

GSC is funded by the National Institute for Health Research (NIHR) Oxford Biomedical Research Centre (BRC) and by Cancer Research UK program grant (C49297/A27294). PD is funded by the NIHR Oxford BRC. The views expressed are those of the authors and not necessarily those of the NHS, NIHR, or Department of Health.

### Author Declarations

Ethical approval is not requiered because only published data is used.

## REFERENCES

1. Moons KGM, Royston P, Vergouwe Y, Grobbee DE, Altman DG. Prognosis and prognostic research: What, why, and how? BMJ. 2009;338(7706):1317–1320. doi:10.1136/bmj.b375

2. Steyerberg EW, Moons KGM, van der Windt DA, et al. Prognosis Research Strategy (PROGRESS) 3: Prognostic Model Research. PLoS Med. 2013;10(2). doi:10.1371/journal.pmed.1001381

3. Riley, Richard D; van der Windt, Danielle; Croft, Peter; Moons KGM. Prognosis Research in Health Care: Concepts, Methods, and Impact. Oxford University Press; 2019. doi:10.1093/med/9780198796619.001.0001

4. Damen Jaag, Hooft L, Schuit E, et al. Prediction models for cardiovascular disease risk in the general population: Systematic review. BMJ. 2016;353. doi:10.1136/bmj.i2416

5. Bi Q, Goodman KE, Kaminsky J, Lessler J. What is machine learning? A primer for the epidemiologist. Am J Epidemiol. 2019;188(12):2222–2239. doi:10.1093/aje/kwz189

6. Mitchell T. Machine Learning. McGraw Hill; 1997.

7. Heus P, Damen Jaag, Pajouheshnia R, et al. Poor reporting of multivariable prediction model studies: Towards a targeted implementation strategy of the TRIPOD statement. BMC Med. 2018;16(1):1–12. doi:10.1186/s12916-018-1099-2

8. Bouwmeester W, Zuithoff NPA, Mallett S, et al. Reporting and methods in clinical prediction research: A systematic review. PLoS Med. 2012;9(5). doi:10.1371/journal.pmed.1001221

9. Collins GS, Mallett S, Omar O, Yu LM. Developing risk prediction models for type 2 diabetes: A systematic review of methodology and reporting. BMC Med. 2011;9. doi:10.1186/1741-7015-9-103

10. Collins GS, De Groot JA, Dutton S, et al. External validation of multivariable prediction models: A systematic review of methodological conduct and reporting. BMC Med Res Methodol. 2014;14(1):40. doi:10.1186/1471-2288-14-40

11. Zamanipoor Najafabadi AH, Ramspek CL, Dekker FW, et al. TRIPOD statement: a preliminary pre-post analysis of reporting and methods of prediction models. BMJ Open. 2020;10(9):e041537. doi:10.1136/bmjopen-2020-041537

12. Christodoulou E, Ma J, Collins GS, Steyerberg EW, Verbakel JY, Van Calster B. A systematic review shows no performance benefit of machine learning over logistic regression for clinical prediction models. J Clin Epidemiol. 2019;110:12–22. doi:10.1016/j.jclinepi.2019.02.004

13. Gravesteijn BY, Nieboer D, Ercole A, et al. Machine learning algorithms performed no better than regression models for prognostication in traumatic brain injury. J Clin Epidemiol. 2020;122:95–107. doi:10.1016/j.jclinepi.2020.03.005

14. Glasziou P, Altman DG, Bossuyt P, et al. Reducing waste from incomplete or unusable reports of biomedical research. Lancet. 2014;383(9913):267–276. doi:10.1016/S0140-6736(13)62228-X

15. Moons KGM, Altman DG, Reitsma JB, et al. Transparent reporting of a multivariable prediction model for individual prognosis or diagnosis (TRIPOD): Explanation and elaboration. Ann Intern Med. 2015;162(1):W1–W73. doi:10.7326/M14-0698

16. Collins GS, Reitsma JB, Altman DG, Moons KGM. Transparent Reporting of a multivariable prediction model for Individual Prognosis Or Diagnosis (TRIPOD): The TRIPOD Statement. Ann Intern Med. 2015;162(1):55. doi:10.7326/M14-0697

17. Collins GS, M Moons KG. Reporting of artificial intelligence prediction models. Published online 2019. doi:10.1016/S0140-6736(19)30235-1

18. Andaur Navarro CL, Damen Jaag, Takada T, et al. Protocol for a systematic review on the methodological and reporting quality of prediction model studies using machine learning techniques. BMJ Open. 2020;10(11):1–6. doi:10.1136/bmjopen-2020-038832

19. Moher D, Liberati A, Tetzlaff J, et al. Preferred reporting items for systematic reviews and meta-analyses: The PRISMA statement. PLoS Med. 2009;6(7). doi:10.1371/journal.pmed.1000097

20. Ouzzani M, Hammady H, Fedorowicz Z, Elmagarmid A. Rayyan-a web and mobile app for systematic reviews. Syst Rev. 2016;5(1):210. doi:10.1186/s13643-016-0384-4

21. Heus P, Damen Jaag, Pajouheshnia R, et al. Uniformity in measuring adherence to reporting guidelines: The example of TRIPOD for assessing completeness of reporting of prediction model studies. BMJ Open. 2019;9(4). doi:10.1136/bmjopen-2018-025611

22. Harris PA, Taylor R, Minor BL, et al. The REDCap consortium: Building an international community of software platform partners. J Biomed Inform. 2019;95:103208. doi:10.1016/j.jbi.2019.103208

23. Chen T, Guestrin C. XGBoost: A scalable tree boosting system. In: Proceedings of the ACM SIGKDD International Conference on Knowledge Discovery and Data Mining. Vol 13-17-August-2016. Association for Computing Machinery; 2016:785–794. doi:10.1145/2939672.2939785

24. Therneau TM, Atkinson EJ. An Introduction to Recursive Partitioning Using the RPART Routines.; 1997.

25. Austin PC, Steyerberg EW. Events per variable (EPV) and the relative performance of different strategies for estimating the out-of-sample validity of logistic regression models. Stat Methods Med Res. 2017;26(2):796–808. doi:10.1177/0962280214558972

26. Yusuf M, Atal I, Li J, et al. Reporting quality of studies using machine learning models for medical diagnosis: a systematic review. BMJ Open. 2020;10(3):e034568. doi:10.1136/bmjopen-2019-034568

27. Nagendran M, Chen Y, Lovejoy CA, et al. Artificial intelligence versus clinicians: Systematic review of design, reporting standards, and claims of deep learning studies in medical imaging. BMJ. 2020;368. doi:10.1136/bmj.m689

28. Faes L, Liu X, Wagner SK, et al. A clinician’s guide to artificial intelligence: How to critically appraise machine learning studies. Transl Vis Sci Technol. 2020;9(2):7–7. doi:10.1167/tvst.9.2.7

29. Liu X, Faes L, Kale AU, et al. A comparison of deep learning performance against health-care professionals in detecting diseases from medical imaging: a systematic review and meta-analysis. Lancet Digit Heal. 2019;1(6):e271–e297. doi:10.1016/S2589-7500(19)30123-2

30. Simera I, Moher D, Hirst A, Hoey J, Schulz KF, Altman DG. Transparent and accurate reporting increases reliability, utility, and impact of your research: Reporting guidelines and the EQUATOR Network. BMC Med. 2010;8(1):24. doi:10.1186/1741-7015-8-24

31. Luo W, Phung D, Tran T, et al. Guidelines for developing and reporting machine learning predictive models in biomedical research: A multidisciplinary view. J Med Internet Res. 2016;18(12). doi:10.2196/jmir.5870

32. Norgeot B, Quer G, Beaulieu-Jones BK, et al. Minimum information about clinical artificial intelligence modeling: the MI-CLAIM checklist. Nat Med. 2020;26(9):1320–1324. doi:10.1038/s41591-020-1041-y

33. Moher D, Schulz KF, Simera I, Altman DG. Guidance for developers of health research reporting guidelines. PLoS Med. 2010;7(2). doi:10.1371/journal.pmed.1000217

