## supplemental material for "Completeness of reporting of clinical prediction models developed using supervised machine learning: A systematic review"

Contents

Section 1: Search strategy…………………………………………………………………………………..………...2

Section 2: Definitions…………………………………………………………………….…………………………..….4

Section 3: Characteristics of included studies …..………………………………….………………………5

**Section 1: Search strategy**

Search date: 19 November 2019

1. Machine Learning[MeSH Terms]
2. Deep learning[MeSH Terms]
3. supervised machine learning[MeSH Terms]
4. "Neural Networks, Computer"[Mesh]
5. data mining[MeSH Terms]
6. machine[tiab] AND (learn* OR model*)
7. (statistical[tiab] OR "statistical to learning"[tiab]) AND ( strateg*[tiab])
8. multilayer perceptron*[tiab] OR random forest*[tiab] OR bayes* network*[tiab] OR support vector machine*[tiab] OR nearest neighbor*[tiab] OR k nearest neighbor*[tiab] OR elastic net[tiab] OR naive bayes*[tiab]
9. (classification[tiab] OR regression[tiab] OR estimation[tiab] OR decision[tiab]) AND tree[tiab]
10. ridge[tiab] OR kernel[tiab] OR ensemble[tiab] OR bagging[tiab] OR bagged[tiab] OR boosting[tiab] OR boosted[tiab] OR fuzzy[tiab]
11. #1 OR #2 OR #3 OR #4 OR #5 OR #6 OR #7 OR #8 OR #9 OR #10
12. (Validat* OR Predict* OR Rule*). [tiab]
13. (Predict* AND (Outcome* OR Risk* OR Model*). [tiab]
14. ((History OR Variable* OR Criteria OR Scor* OR Characteristic* OR Finding* OR Factor*) AND (Predict* OR Model* OR Decision* OR Identif* OR Prognos*)). [tiab]
15. (Decision* AND (Model* OR Clinical*). [tiab]
16. (Prognostic AND (History OR Variable* OR Criteria OR Scor* OR Characteristic* OR Finding* OR Factor* OR Model*). [tiab]
17. #12 OR #13 OR #14 OR #15 OR #16
18. (discrimination[tiab] OR discriminative[tiab] OR discriminatory[tiab]) AND (accuracy[tiab] OR ability[tiab] OR performance[tiab] OR value[tiab] OR model[tiab] OR models[tiab] OR power[tiab] OR capacity[tiab] OR capabilit*[tiab] OR efficiency[tiab])
19. (discriminability[tiab] OR c to index[tiab] OR c to statistic[tiab] OR concordance[tiab] OR DCA[tiab])
20. "decision curve"[tiab]
21. calibrat*[tiab] AND (plot*[tiab] OR curve*[tiab] OR slope*[tiab] OR model[tiab] OR models[tiab])
22. performance[tiab] AND (classification[tiab] OR classifier[tiab] OR clinical[tiab] OR accuracy[tiab] OR validation[tiab] OR metrics[tiab] OR diagnostic[tiab] OR AUC[tiab])
23. (sensitivity[tiab] OR specificity[tiab] OR PPV[tiab] OR NPV[tiab])
24. "correctly classified"[tiab]
25. "clinical accuracy"[tiab]
26. positive predictive value*[tiab]
27. negative predictive value*[tiab]
28. classification[tiab] OR classifier[tiab]
29. Area Under Curve[Mesh]
30. "Area under the curve"[tiab]
31. "Area under the ROC"[tiab]
32. “Area Under the Receiver”[tiab]
33. (ROC[tiab] OR AUC[tiab] OR AUROC[tiab])
34. ROC Curve [Mesh]
35. "Hosmer to Lemeshow"[tiab] OR "H to L test"[tiab]
36. "expected ratio"[tiab] OR "observed ratio"[tiab] OR "E:O ratio"[tiab]
37. #18 OR #19 OR #20 OR #21 OR #22 OR #23 OR #24 OR #25 OR #26 OR #27 OR #28 OR #29 OR #30 OR #31 OR #32 OR #33 OR #34 OR #35 OR #36
38. #11 AND #17
39. #11 AND (#17 OR #37)
40. #39 AND (“2018/01/01”[PDat]: “2019/12/31”[PDat])
41. #40 NOT “review”[pt]
42. #39 AND (“2019/01/01”[PDat]: “2019/12/31”[PDat])
43. #42 NOT “review”[pt]

Results #41= **24732**

Results #43=**12977**

**Section 2: Definitions**

*Definition of research aim*

*Model development studies* aim to develop a prediction model to be used for individualized predictions where its predictive performance is directly evaluated using exactly the same data, either directly (apparent performance) or by resampling participant data or random/non to random split of the data (internal validation). *Model development studies with external validation (same model)* also describe the development of a model, but this is followed by quantifying its predictive performance in different participants, i.e. external to the development sample. *Model development studies with external validation (different model)* either aim to develop a prediction model and compare it to the performance of existing models that were developed previously, or to update or adjust an existing model that performs poorly by recalibrating or extending the model. *External validation only* studies aim to assess only the predictive performance of existing prediction models using data external to the development sample. (1,2)

*Definition of performance measures*

Discrimination refers to the ability of a prediction model to differentiate between those who do or do not experience the outcome event. The most common reported measure for discrimination is the concordance index (c to index) and the area under the receiver to operating characteristic curves. Calibration reflects the agreement between predictions made by the model and observed outcomes. Calibration is preferably reported graphically, with observed risk plotted on the y to axis against predicted risk on the x to axis. (1,2)

**References**

1.	Collins GS, Reitsma JB, Altman DG, Moons KGM. Transparent Reporting of a multivariable prediction model for Individual Prognosis Or Diagnosis (TRIPOD): The TRIPOD Statement. Ann Intern Med. 2015 Jan 6;162(1):55.

2.	Moons KGM, Altman DG, Reitsma JB, Ioannidis JPA, Macaskill P, Steyerberg EW, et al. Transparent reporting of a multivariable prediction model for individual prognosis or diagnosis (TRIPOD): Explanation and elaboration. Ann Intern Med. 2015 Jan 6;162(1):W1 to 73.

**Section 3. Characteristics of included studies**

| **Table S1.** Characteristics of the included studies (n=152) | | | | | | | |
| --- | --- | --- | --- | --- | --- | --- | --- |
| **First Author** | **Journal** | **Impact factor** ^a^ | **Publication year** | **Clinical field** | **Study type** | **Study design** | **Outcome** |
| X Jiang^1^ | PLoS ONE | 2.740 | 2019 | Oncology | Prognosis | Development with external validation (same model) | 5-year breast cancer metastasis |
| L Adhikari^2^ | PLoS ONE | 2.740 | 2019 | Nephrology | Prognosis | Development only (including internal validation) | Acute kidney injury at first 7 days after surgery |
| WP Chen^3^ | BioMed Research International | 2.276 | 2018 | Dentistry | Diagnosis | Development only (including internal validation) | Periodontitis |
| G Lorenzoni^4^ | Journal of Clinical Medicine | 3.303 | 2019 | Cardiovascular medicine | Prognosis | Development only (including internal validation) | First hospitalization in heart failure patients |
| L-K Pries^5^ | Schizophrenia Bulletin | 7.958 | 2019 | Psychiatry | Prognosis | Development with external validation (same model) | Schizophrenia |
| I Sánchez Fernández^6^ | Journal of Child Neurology | 2.092 | 2018 | Neurology | Prognosis | Development only (including internal validation) | In-hospital mortality in critically ill children monitored with cEEG in the ICU |
| H Zhang^7^ | GigaScience | 4.688 | 2018 | Neurology | Diagnosis | Development with external validation (same model) | Alzheimer’s disease |
| A Koivu^8^ | Computers in Biology and Medicine | 2.286 | 2018 | Obstetrics & Gynecology | Diagnosis | Development with external validation (same model) | First trimester prenatal down's syndrome |
| GGP Garcia^9^ | American Journal of Ophthalmology | 4.483 | 2018 | Ophthalmology | Prognosis | Development only (including internal validation) | progression normal tension glaucoma |
| K Kajiwara^10^ | Journal of Vascular and Interventional Radiology | 2.828 | 2018 | Oncology | Diagnosis | Development only (including internal validation) | Insulinomas |
| A Tam^11^ | GigaScience | 5.993 | 2019 | Neurology | Prognosis | Development with external validation (same model) | Progression to Alzheimer’s dementia |
| LC Chambers^12^ | Sexually Transmitted Diseases | 2.270 | 2018 | Healthcare services | Diagnosis | Development only (including internal validation) | Need for a standard visit |
| V Bhat^13^ | Mayo Clinic Proceeding | 7.091 | 2018 | Surgery | Prognosis | Development only (including internal validation) | New-onset diabetes after transplant |
| H Won Choi^14^ | American Journal of Roentgenology | 3.161 | 2018 | Medical imaging | Diagnosis | Development only (including internal validation) | Early prediction of the severity of acute pancreatitis |
| A Ogunleye^15^ | IEEE/ACM Transactions on Computational Biology and Bioinformatics(^c^) | 3.015 | 2019 | Nephrology | Diagnosis | Development only (including internal validation) | Chronic kidney disease |
| CC Wu^16^ | Computer Methods and Programs in Biomedicine | 3.424 | 2018 | Hepatology | Diagnosis | Development only (including internal validation) | Early fatty liver disease |
| D Shigemi^17^ | The Journal of Maternal-Fetal & Neonatal Medicine | 1.737 | 2019 | Obstetrics & Gynecology | Diagnosis | Development only (including internal validation) | Macrosomia |
| KG Friedman^18^ | Ultrasound in Obstetrics & Gynecology | 5.595 | 2018 | Neonatology | Prognosis | Development only (including internal validation) | Circulation type |
| H Duan^19^ | BMC Medical Informatics and Decision Making | 2.317 | 2019 | Cardiovascular medicine | Prognosis | Development only (including internal validation) | Major adverse cardiac event |
| M Ansart^20^ | Statistical Methods in Medical Research | 2.291 | 2019 | Neurology | Diagnosis | Development with external validation (same model) | Brain amyloidosis |
| J Kwon^21^ | Resuscitation | 4.215 | 2019 | Cardiovascular medicine | Prognosis | Development only (including internal validation) | neurological recovery after ROSC |
| R Hammond^22^ | PLoS ONE | 2.740 | 2019 | Nutrition | Prognosis | Development only (including internal validation) | Obesity status at the age of five |
| S Zamboni^23^ | World Journal of Urology | 3.217 | 2019 | Oncology | Diagnosis | Development only (including internal validation) | Adverse pathologic features |
| AL Nobles^24^ | Proceedings of the SIGCHI Conference on Human Factor in Computing Systems(^c^) | - | 2018 | Psychiatry | Prognosis | Development only (including internal validation) | Suicidality |
| R Tse^25^ | American Journal of Forensic Medicine and Pathology | 0.539 | 2018 | Forensic pathology | Diagnosis | Development only (including internal validation) | Salt water drowning with immersion time of less than 1 hour (SWD1) |
| NW Sterling^26^ | International Journal of Medical Informatics | 3.025 | 2019 | Emergency medicine | Prognosis | Development only (including internal validation) | ED disposition |
| S Perveen^27^ | Scientific Reports | 4.011 | 2018 | Hepatology | Diagnosis | Development only (including internal validation) | Non-alcoholic fatty liver disease risk |
| Z Pei^28^ | Interdisciplinary Sciences-Computational life Sciences | 1.418 | 2018 | Primary care | Diagnosis | Development only (including internal validation) | Essential hypertension |
| FB Bouallegue^29^ | Journal of Alzheimer's Disease | 3.517 | 2018 | Neurology | Prognosis | Development only (including internal validation) | Alzheimer's disease |
| T-L Tsai^30^ | Journal of Clinical Medicine | 3.303 | 2019 | Critical care | Prognosis | Development only (including internal validation) | Successful extubation |
| RR Lopes^31^ | Netherlands Heart Journal | 1.933 | 2019 | Cardiovascular medicine | Prognosis | Development only (including internal validation) | Mortality |
| G Maragatham^32^ | Journal of Medical Systems | 3.058 | 2019 | Cardiovascular medicine | Prognosis | Development only (including internal validation) | Heart failure |
| C-Y Shao^33^ | Thoracic Cancer | 2.610 | 2019 | Surgery | Prognosis | Development only (including internal validation) | anastomosis leakage after esophagectomy |
| V Sacca^34^ | Brain Imaging and Behavior | 3.418 | 2018 | Neurology | Diagnosis | Development only (including internal validation) | Early multiple sclerosis |
| M Cearns^35^ | Translational Psychiatry | 5.280 | 2019 | Psychiatry | Prognosis | Development only (including internal validation) | Re-hospitalization within 2 years of major depressive episode |
| MJRJ Bouts^36^ | Human Brain Mapping | 4.421 | 2019 | Neurology | Diagnosis | Development with external validation (same model) | Mild cognitive impairment |
| NB Huben^37^ | Journal of Endourology | 2.267 | 2018 | Urology | Prognosis | Development only (including internal validation) | Operative time for RARP |
| AT Hale^38^ | Neurosurgical focus | 2.891 | 2018 | Critical care | Prognosis | Development only (including internal validation) | Death or alive with GOS score ≤ 3 |
| C Salvatore^39^ | Journal of Neuroscience Methods | 2.785 | 2018 | Neurology | Prognosis | Development only (including internal validation) | Cognitive status (HC; ncMCI; cMCI; AD) |
| H Yang^40^ | IEEE Journal of Biomedical and Health Informatics | 5.223 | 2019 | Neurology | Diagnosis | Development only (including internal validation) | Dementia |
| M Zhou^41^ | BMC Medical Informatics and Decision Making | 2.317 | 2019 | Preventive care | Prognosis | Development only (including internal validation) | Exercise relapse |
| X Kang^42^ | Journal of Maternal-Fetal & Neonatal Medicine | 1.737 | 2019 | Obstetrics & Gynecology | Prognosis | Development only (including internal validation) | Gestational diabetes mellitus with macrosomia |
| CM Sauer^43^ | PLoS ONE | 2.776 | 2018 | Infectious diseases | Prognosis | Development only (including internal validation) | Tuberculosis treatment failure |
| LW Thornblade^44^ | Journal for Electronic Health data and Methods(^c^) | - | 2018 | Surgery | Prognosis | Development only (including internal validation) | Elective colon resection |
| VJ Lei^45^ | Studies in Health Technology and Informatics(^b^) | 0.71 | 2019 | Surgery | Prognosis | Development only (including internal validation) | All-cause in-hospital mortality |
| SJ Lee^46^ | Studies in Health Technology and Informatics(^b^) | 0.71 | 2019 | Oncology | Prognosis | Development only (including internal validation) | Cancer recurrence |
| GB Auffenberg^47^ | European Urology | 18.728 | 2019 | Urology | Prognosis | Development only (including internal validation) | Prostate cancer treatment option |
| Z Wang^48^ | Journal of Biomedical Informatics | 3.526 | 2019 | Cardiovascular medicine | Prognosis | Development only (including internal validation) | 1-year mortality |
| A Nelson^49^ | np Digital Medicine | 0.00 | 2019 | Healthcare services | Prognosis | Development only (including internal validation) | Schedule appointment attendance |
| T Shibahara^50^ | JCO Clinical Cancer Informatics(^b^) | 0.43 | 2018 | Oncology | Prognosis | Development only (including internal validation) | Blood cell count |
| S Liang^51^ | Schizophrenia Research | 4.569 | 2018 | Psychiatry | Diagnosis | Development only (including internal validation) | Schizophrenia/depression/  Healthy/controls |
| R Chen^52^ | Circulation-Cardiovascular Quality and Outcomes | 5.071 | 2019 | Cardiovascular medicine | Prognosis | Development only (including internal validation) | Heart failure |
| E Klang^53^ | Neuroradiology | 2.238 | 2019 | Medical imaging | Diagnosis | Development only (including internal validation) | Use of non-contrast CT in ED department |
| Y Fan^54^ | Endocrine | 3.235 | 2019 | Surgery | Prognosis | Development only (including internal validation) | Tumor remission after transphenoidal surgery (TSS) |
| A Ferre^55^ | Journal of Clinical Sleep Medicine | 3.586 | 2019 | Neurology | Prognosis | Development with external validation (same model) | RDI equal to or above 10 events/h |
| F Zhang^56^ | Metabolomics | 3.167 | 2018 | Oncology | Prognosis | Development only (including internal validation) | Recurrence of Epithelial Ovarian Cancer at 5-years |
| R Ferrer-Peña^57^ | Journal of Manipulative and Physiological Therapeutics | 1.230 | 2019 | Physical medicine | Diagnosis | Development only (including internal validation) | Needle length |
| JM Cameron^58^ | Analyst | 3.978 | 2019 | Oncology | Diagnosis | Development only (including internal validation) | Brain tumor |
| AHS Harris^59^ | The Journal of Arthroplasty | 3.524 | 2018 | Surgery | Prognosis | Development only (including internal validation) | 30-day mortality |
| KM Kuo^60^ | BMC Medical Informatics and Decision Making | 2.317 | 2019 | Psychiatry | Prognosis | Development only (including internal validation) | Hospital-acquired pneumonia |
| Y Arai^61^ | Blood advances | 4.910 | 2019 | Immunology | Prognosis | Development only (including internal validation) | Acute graft-versus-host disease |
| JK Paul^62^ | Computers in Biology and Medicine | 3.434 | 2019 | Neurology | Diagnosis | Development only (including internal validation) | Fibromyalgia |
| L Liu^63^ | BMC Systems Biology | 2.048 | 2018 | Traumatology | Prognosis | Development only (including internal validation) | Side effects of analgesics |
| C Shappell^64^ | Critical Care Medicine | 6.971 | 2018 | Critical care | Prognosis | Development only (including internal validation) | In-hospital mortality |
| X Niu^65^ | Scientific Reports | 4.011 | 2018 | Cardiovascular medicine | Prognosis | Development only (including internal validation) | MACEs within 1-year follow-up |
| F Ge^66^ | Journal of Affective Disorders | 3.892 | 2019 | Psychiatry | Prognosis | Development only (including internal validation) | Posttraumatic stress disorder at 3 months |
| B Dhondt^67^ | World Journal of Urology | 3.217 | 2019 | Oncology | Diagnosis | Development only (including internal validation) | Pligometastic vs polymetastatic in prostatic cancer |
| B Rohaut^68^ | Scientific Reports | 3.998 | 2019 | Medical imaging | Prognosis | Development only (including internal validation) | Consciousness at ICU discharge |
| JM Karnuta^69^ | The Journal of Arthroplasty | 3.524 | 2019 | Surgery | Prognosis | Development only (including internal validation) | Inpatient payments prior to lower extremity arthroplasty |
| MB Wilson^70^ | Otolaryngology-Head and Neck Surgery | 2.341 | 2019 | Otolaryngology | Diagnosis | Development only (including internal validation) | Peritonsillar abscess |
| B Lu^71^ | Sensors | 3.275 | 2019 | Oncology | Diagnosis | Development only (including internal validation) | Lung cancer |
| Y Xu^72^ | BMC Cancer | 3.150 | 2019 | Oncology | Diagnosis | Development only (including internal validation) | Breast cancer recurrence |
| S Cohen^73^ | Autism Research | 3.697 | 2018 | Psychiatry | Prognosis | Development only (including internal validation) | Daytime challenging behaviors |
| Y Wang^74^ | Academic Radiology | 2.488 | 2019 | Medical imaging | Diagnosis | Development only (including internal validation) | Differentiation between T2 and T3/T4 stage in gastric cancer |
| A Mortezagholi^75^ | Asian pacific journal of cancer prevention | 0.00 | 2019 | Oncology | Diagnosis | Development only (including internal validation) | Gastric cancer |
| UJ Muehlematter^76^ | European Radiology | 3.962 | 2018 | Medical imaging | Diagnosis | Development only (including internal validation) | Vertebral insufficiency fractures |
| SHA Faruqui^77^ | JMIR MHealth and UHealth | 4.313 | 2019 | Endocrinology | Prognosis | Development only (including internal validation) | Blood glucose level for type 2 Diabetes Mellitus |
| M Molinari^78^ | Transplantation | 4.546 | 2019 | Surgery | Prognosis | Development only (including internal validation) | 90-day mortality |
| JP Jeon^79^ | Clinical Neurology and Neurosurgery | 1.672 | 2018 | Surgery | Diagnosis | Development only (including internal validation) | Persistent hemodynamic depression following CAS |
| VE Staartjes^80^ | Neurosurgical Focus | 2.891 | 2018 | Surgery | Prognosis | Development only (including internal validation) | Gross-total resection in transspheinoidal surgery for pituitary adenoma at 3 months |
| C-F Lu^81^ | Clinical Cancer Research | 8.911 | 2018 | Medical imaging | Diagnosis | Development with external validation (same model) | Glioblastoma vs lower grade gliomas |
| N Park^82^ | PLoS ONE | 2.776 | 2018 | Oncology | Prognosis | Development only (including internal validation) | AKI occurrence in 14 days |
| T Ballarini^83^ | NeuroImage: Clinical 21 | 4.350 | 2019 | Neurology | Diagnosis | Development only (including internal validation) | Individual treatment response |
| D Chen^84^ | Clinical Cancer Informatics(^b^) | 0.43 | 2019 | Oncology | Prognosis | Development only (including internal validation) | Time to first treatment in Chronic Lymphocytic Leukemia |
| J Malycha^85^ | Resuscitation | 4.215 | 2019 | Critical care | Prognosis | Development only (including internal validation) | FiO2 Added value |
| Z Xie^86^ | Preventing chronic disease | 2.144 | 2019 | Endocrinology | Prognosis | Development only (including internal validation) | type 2 diabetes risk |
| B Thanathornwong^87^ | Health Informatics Research | 2.939 | 2018 | Dentistry | Diagnosis | Development only (including internal validation) | Need of orthodontic treatment in permanent dentition |
| CQ Ngo^88^ | Annual International Conference of the IEEE Engineering in Medicine and Biology Society(^b^) | 1.01 | 2018 | Endocrinology | Diagnosis | Development only (including internal validation) | Hypoglycemia episode |
| SH Hyun^89^ | Clinical Nuclear Medicine | 6.622 | 2019 | Oncology | Diagnosis | Development only (including internal validation) | adenocarcinoma vs squamous cell carcinoma |
| A Rozet^90^ | Journal of Medical Internet Research | 5.034 | 2019 | Psychiatry | Prognosis | Development only (including internal validation) | Self-reported stress over 100 days |
| MS Mellem^91^ | Biological Psychiatry: CNNI | 5.335 | 2019 | Psychiatry | Diagnosis | Development only (including internal validation) | Transdiagnostic Symptom Severity |
| AV Karhade^92^ | The Spine Journal | 3.191 | 2019 | Surgery | Prognosis | Development only (including internal validation) | Prolonged opioid prescription after surgery for lumbar disc herniation to at least 90 to 180 days postoperatively |
| F Zhang^93^ | Neuroscience | 5.679 | 2019 | Neurology | Diagnosis | Development only (including internal validation) | Alzheimer's disease |
| D Leightley^94^ | Journal of Mental Health | 2.604 | 2018 | Psychiatry | Diagnosis | Development only (including internal validation) | Post-traumatic stress disorder |
| M Mulder^95^ | Archives of Physical Medicine and Rehabilitation | 3.098 | 2019 | Neurology | Prognosis | Development only (including internal validation) | Community walkers after stoke |
| J Debedat^96^ | Diabetes Care | 15.270 | 2018 | Endocrinology | Prognosis | Development with external validation (same model) | Type 2 diabetes relapse after Gastric Bypass |
| JCR Alcantud^97^ | PLoS ONE | 2.740 | 2019 | Oncology | Prognosis | Development only (including internal validation) | 5-years survival rate |
| A Sandstrom^98^ | PLoS ONE | 2.740 | 2019 | Obstetrics & Gynecology | Diagnosis | Development only (including internal validation) | Preeclampsia with delivery <34 weeks of gestation |
| C Xiao^99^ | Annual International Conference of the IEEE Engineering in Medicine and Biology Society(^b^) | 1.01 | 2018 | Neurology | Diagnosis | Development only (including internal validation) | Parkinson's disease |
| JN Cooper^100^ | Journal of surgical research | 1.872 | 2018 | Surgery | Prognosis | Development with external validation (same model) | 30-day postoperative neonatal mortality |
| C-S Rau^101^ | PLoS ONE | 2.776 | 2018 | Surgery | Prognosis | Development only (including internal validation) | In-hospital mortality after severe traumatic brain injury |
| RS Anand^102^ | AMIA Joint Summits on Translational Sciences Proceedings(^c^) | - | 2018 | Critical care | Prognosis | Development only (including internal validation) | All cause in-hospital mortality |
| Y Aperstein^103^ | PLoS ONE | 2.740 | 2019 | Critical care | Prognosis | Development only (including internal validation) | ICU mortality |
| J Park^104^ | Journal of Medical Internet Research | 5.034 | 2019 | Cardiovascular medicine | Prognosis | Development with external validation (same model) | Cardio-cerebrovascular event in patients with hypertension |
| D Gökçay^105^ | IEEE Journal of Biomedical and Health Informatics | 5.223 | 2019 | Rheumatology | Diagnosis | Development only (including internal validation) | Fibromyalgia |
| R Gupta^106^ | Canadian Journal of Ophthalmology | 1.369 | 2019 | Ophthalmology | Prognosis | Development only (including internal validation) | Visual outcome after open globe injury |
| AH Butt^107^ | BioMedical Engineering OnLine | 2.013 | 2018 | Neurology | Diagnosis | Development only (including internal validation) | Patients with Parkinson disease |
| HS Hunter-Zinck^108^ | Journal of the American Medical Informatics Association | 4.112 | 2019 | Healthcare services | Diagnosis | Development only (including internal validation) | Emergency department orders |
| A Kilic^109^ | Annals of Thoracic Surgery | 3.639 | 2019 | Surgery | Prognosis | Development only (including internal validation) | Operative mortality |
| C Campillo-Artero^110^ | PLoS One | 2.776 | 2018 | Obstetrics & Gynaecology | Prognosis | Development only (including internal validation) | Emergency cesarean section |
| ML Zhang^111^ | American Journal of Clinical Pathology | 2.094 | 2019 | Pathology | Diagnosis | Development only (including internal validation) | PBFC with current/recent CBC/differential |
| WS Hong^112^ | PLoS One | 2.776 | 2018 | Healthcare services | Prognosis | Development only (including internal validation) | patient's disposition (discharge, admission) |
| O Beauchet^113^ | Journal of Nutrition Health and Aging | 2.660 | 2018 | Geriatric | Prognosis | Development only (including internal validation) | Fall in acute care medical wards |
| Z Ma^114^ | PLoS One | 2.776 | 2018 | Cardiovascular medicine | Prognosis | Development only (including internal validation) | Warfarin dose |
| SPK Veeranki^115^ | Studies in Health Technology and Informatics(^b^) | 0.71 | 2019 | Neurology | Prognosis | Development only (including internal validation) | Delirium |
| H Maharlou^116^ | Healthcare Informatics Research | 2.939 | 2018 | Healthcare services | Prognosis | Development only (including internal validation) | Length of stay in ICU after cardiac surgery |
| C Castillo-Olea^117^ | International Journal of Environmental Research and Public Health | 2.468 | 2019 | Geriatric | Diagnosis | Development only (including internal validation) | Sarcopenia |
| C Sa-ngamuang^118^ | PLoS neglected tropical diseases | 4.487 | 2018 | Infectious diseases | Diagnosis | Development only (including internal validation) | Dengue |
| A Talaei-Khoei^119^ | International Journal of Medical Informatics | 2.731 | 2018 | Endocrinology | Prognosis | Development only (including internal validation) | Type 2 diabetes risk at 1, 3 and 8 years |
| D Bertsimas^120^ | Annals of Surgery | 9.476 | 2018 | Surgery | Prognosis | Development with external validation (same model) | 30-day mortality |
| C Liu^121^ | Abdominal Radiology | 2.429 | 2019 | Oncology | Prognosis | Development only (including internal validation) | Lymphadenectomy extension in gastric cancer before surgical resection |
| G Luo^122^ | JMIR Medical Informatics | 2.577 | 2019 | Healthcare services | Prognosis | Development with external validation (same model) | Appropriate hospital admission for patients with bronchiolitis |
| K Meena^123^ | Artificial Intelligence in Medicine | 4.383 | 2019 | Pediatrics | Diagnosis | Development only (including internal validation) | Anemia status in children |
| M TakeuchI^124^ | Journal of Gastrointestinal Surgery | 2.686 | 2018 | Oncology | Prognosis | Development only (including internal validation) | Post-operative overall survival and disease-free survival |
| H Kiiski^125^ | Brain Topography | 3.104 | 2018 | Neurology | Prognosis | Development only (including internal validation) | Cognitive functioning and processing speed over 2-year |
| Z Hasnain^126^ | PLoS ONE | 2.740 | 2019 | Oncology | Prognosis | Development only (including internal validation) | Post-cystectomy recurrence |
| J Dean^127^ | Clinical and Translational Radiation Oncology | 1.439 | 2018 | Oncology | Prognosis | Development with external validation (same model) | Severe acute dysphagia resulting from head and neck radiotherapy |
| M Cheung^128^ | Surgery | 3.476 | 2018 | Surgery | Prognosis | Development only (including internal validation) | Mortality in burn patients |
| JL Gowin^129^ | NeuroImage: Clinical | 4.350 | 2019 | Psychiatry | Prognosis | Development with external validation (same model) | Relapse rate at 12 months after treatment |
| JC Rojas^130^ | Annals of the American Thoracic Society | 4.026 | 2018 | Healthcare services | Prognosis | Development with external validation (same model) | Intensive care unit readmission |
| R Wei^131^ | Technology in Cancer Research & Treatment | 2.074 | 2019 | Medical imaging | Diagnosis | Development with external validation (same model) | Pre-operative serous cystic neoplasms |
| J Balani^132^ | Obstetric Medicine(^b^) | 0.389 | 2018 | Endocrinology | Prognosis | Development only (including internal validation) | Gestational diabetes mellitus |
| L Gao^133^ | Journal of Neurotrauma | 4.056 | 2019 | Critical care | Prognosis | Development only (including internal validation) | Mortality after severe traumatic brain injury at 6 months |
| A Garcia-Arce^134^ | Journal for Healthcare Quality | 1.092 | 2018 | Healthcare services | Prognosis | Development only (including internal validation) | Preventable readmission within 30-days |
| S Papini^135^ | Journal of Anxiety Disorders | 3.472 | 2018 | Psychiatry | Diagnosis | Development only (including internal validation) | Posttraumatic stress disorders screening status 3 months post hospitalization |
| B-S Jang^136^ | Scientific Reports | 4.011 | 2018 | Medical imaging | Diagnosis | Development with external validation (same model) | Pseudoprogression in patients with glioblastoma |
| CV Cosgriff^137^ | npj Digital Medicine(^c^) | - | 2019 | Critical care | Prognosis | Development only (including internal validation) | Illness severity score |
| H Kim^138^ | JMIR MHealth and UHealth | 4.313 | 2019 | Psychiatry | Diagnosis | Development only (including internal validation) | Depression |
| J Lotsch^139^ | Breast Cancer Research and Treatment | 3.471 | 2018 | Oncology | Prognosis | Development only (including internal validation) | Persistent pain after breast cancer surgery at 3 years |
| II Spyroglou^140^ | BMC Research Notes(^b^) | 1.38 | 2018 | Immunology | Prognosis | Development only (including internal validation) | Asthma exacerbation |
| B Goudey^141^ | Scientific Reports | 3.998 | 2019 | Neurology | Diagnosis | Development only (including internal validation) | Abnormal CSF Aβ1-42 level |
| A Facciorusso^142^ | Pancreatology | 3.629 | 2019 | Oncology | Prognosis | Development only (including internal validation) | Pain response to repeat echoendoscopic celiac plexus neurolysis |
| DW Kim^143^ | Bone | 4.360 | 2018 | Dentistry | Prognosis | Development only (including internal validation) | Occurrence of BRONJ associated with dental extraction |
| AV Karhade^144^ | Spine Journal | 3.191 | 2019 | Surgery | Prognosis | Development only (including internal validation) | In-hospital and 90-day post-discharge mortality in SEA |
| W Tu^145^ | Journal of NeuroVirology | 2.354 | 2019 | Neurology | Diagnosis | Development only (including internal validation) | HIV-associated neurocognitive disorder |
| T van Steenkiste^146^ | Artificial Intelligence in Medicine | 4.383 | 2019 | Critical care | Prognosis | Development only (including internal validation) | Positive blood culture at 72hr |
| N Paliwal^147^ | Neurosurgical Focus | 2.891 | 2018 | Surgery | Prognosis | Development only (including internal validation) | Diverters treatment outcome (Occlusion vs. residual) |
| M Bronsert^148^ | American Journal of Surgery | 2.125 | 2019 | Surgery | Diagnosis | Development only (including internal validation) | Postoperative complications |
| A Eill^149^ | Brain Connectivity | 5.263 | 2019 | Neurology | Diagnosis | Development only (including internal validation) | Autism spectrum disorders |
| F Cook^150^ | British Journal of Anaesthesia | 6.880 | 2019 | Surgery | Diagnosis | Development only (including internal validation) | Intubation difficulty |
| B Eggleston^151^ | Brain Injury | 1.690 | 2019 | Healthcare services | Diagnosis | Development only (including internal validation) | Service-connected disability (SCD) ≥50 among a cohort of veterans with previous combat deployment |
| YR Villarreal^152^ | Social Work in Public Health | 0.607 | 2019 | Primary care | Diagnosis | Development only (including internal validation) | Hepatitis C Virus Incidence |
| ^a^ Value is based on the Journal Citation Report from the year of publication of the article.  ^b^ Value is based on the Scientific Journal Ranking from the year of publication of the article.  ^c^ Value is unavailable. | | | | | | | |

**References**

1.	Jiang X, Wells A, Brufsky A, Neapolitan R. A clinical decision support system learned from data to personalize treatment recommendations towards preventing breast cancer metastasis. *PLoS One*. 2019;14(3):1-18. doi:10.1371/journal.pone.0213292

2.	Adhikari L, Ozrazgat-Baslanti T, Ruppert M, et al. Improved predictive models for acute kidney injury with IDEA: Intraoperative data embedded analytics. *PLoS One*. 2019;14(4):1-26. doi:10.1371/journal.pone.0214904

3.	Chen WP, Chang SH, Tang CY, Liou ML, Tsai SJJ, Lin YL. Composition Analysis and Feature Selection of the Oral Microbiota Associated with Periodontal Disease. *Biomed Res Int*. 2018;2018. doi:10.1155/2018/3130607

4.	Lorenzoni G, Sabato SS, Lanera C, et al. Comparison of Machine Learning Techniques for Prediction of Hospitalization in Heart Failure Patients. *J Clin Med*. 2019;8(9):1298. doi:10.3390/jcm8091298

5.	Pries LK, Lage-Castellanos A, Delespaul P, et al. Estimating Exposome Score for Schizophrenia Using Predictive Modeling Approach in Two Independent Samples: The Results from the EUGEI Study. *Schizophr Bull*. 2019;45(5):960-965. doi:10.1093/schbul/sbz054

6.	Sánchez Fernández I, Sansevere AJ, Gaínza-Lein M, Kapur K, Loddenkemper T. Machine Learning for Outcome Prediction in Electroencephalograph (EEG)-Monitored Children in the Intensive Care Unit. *J Child Neurol*. 2018;33(8):546-553. doi:10.1177/0883073818773230

7.	Zhang H, Zhu F, Dodge HH, Higgins GA, Omenn GS, Guan Y. A similarity-based approach to leverage multi-cohort medical data on the diagnosis and prognosis of Alzheimer’s disease. *Gigascience*. 2018;7(7):1-10. doi:10.1093/gigascience/giy085

8.	Koivu A, Korpimäki T, Kivelä P, Pahikkala T, Sairanen M. Evaluation of machine learning algorithms for improved risk assessment for Down’s syndrome. *Comput Biol Med*. 2018;98(April):1-7. doi:10.1016/j.compbiomed.2018.05.004

9.	Garcia GGP, Nitta K, Lavieri MS, et al. Using Kalman Filtering to Forecast Disease Trajectory for Patients With Normal Tension Glaucoma. *Am J Ophthalmol*. 2019;199:111-119. doi:10.1016/j.ajo.2018.10.012

10.	Kajiwara K, Yamagami T, Toyota N, et al. New Diagnostic Criteria for the Localization of Insulinomas with the Selective Arterial Calcium Injection Test: Decision Tree Analysis. *J Vasc Interv Radiol*. 2018;29(12):1749-1753. doi:10.1016/j.jvir.2018.05.015

11.	Tam A, Dansereau C, Iturria-Medina Y, et al. A highly predictive signature of cognition and brain atrophy for progression to Alzheimer’s dementia. *Gigascience*. 2019;8(5):1-16. doi:10.1093/gigascience/giz055

12.	Chambers LC, Manhart LE, Katz DA, et al. Comparison of Algorithms to Triage Patients to Express Care in a Sexually Transmitted Disease Clinic. *Sex Transm Dis*. 2018;45(10):696-702. doi:10.1097/OLQ.0000000000000854

13.	Bhat V, Tazari M, Watt KD, Bhat M. New-Onset Diabetes and Preexisting Diabetes Are Associated With Comparable Reduction in Long-Term Survival After Liver Transplant: A Machine Learning Approach. *Mayo Clin Proc*. 2018;93(12):1794-1802. doi:10.1016/j.mayocp.2018.06.020

14.	Choi HW, Park HJ, Choi SY, et al. Early prediction of the severity of acute pancreatitis using radiologic and clinical scoring systems with classification tree analysis. *Am J Roentgenol*. 2018;211(5):1035-1043. doi:10.2214/AJR.18.19545

15.	Ogunleye A, Wang QG. XGBoost Model for Chronic Kidney Disease Diagnosis. *IEEE/ACM Trans Comput Biol Bioinforma*. 2020;17(6):2131-2140. doi:10.1109/TCBB.2019.2911071

16.	Wu CC, Yeh WC, Hsu WD, et al. Prediction of fatty liver disease using machine learning algorithms. *Comput Methods Programs Biomed*. 2019;170:23-29. doi:10.1016/j.cmpb.2018.12.032

17.	Shigemi D, Yamaguchi S, Aso S, Yasunaga H. Predictive model for macrosomia using maternal parameters without sonography information. *J Matern Neonatal Med*. 2019;32(22):3859-3863. doi:10.1080/14767058.2018.1484090

18.	Friedman KG, Sleeper LA, Freud LR, et al. Improved technical success, postnatal outcome and refined predictors of outcome for fetal aortic valvuloplasty. *Ultrasound Obstet Gynecol*. 2018;52(2):212-220. doi:10.1002/uog.17530

19.	Duan H, Sun Z, Dong W, Huang Z. Utilizing dynamic treatment information for MACE prediction of acute coronary syndrome. *BMC Med Inform Decis Mak*. 2019;19(1):1-11. doi:10.1186/s12911-018-0730-7

20.	Ansart M, Epelbaum S, Gagliardi G, et al. Reduction of recruitment costs in preclinical AD trials: validation of automatic pre-screening algorithm for brain amyloidosis. *Stat Methods Med Res*. 2020;29(1):151-164. doi:10.1177/0962280218823036

21.	Kwon J myoung, Jeon KH, Kim HM, et al. Deep-learning-based out-of-hospital cardiac arrest prognostic system to predict clinical outcomes. *Resuscitation*. 2019;139(April 2019):84-91. doi:10.1016/j.resuscitation.2019.04.007

22.	Hammond R, Athanasiadou R, Curado S, et al. Correction: Predicting childhood obesity using electronic health records and publicly available data. *PLoS One*. 2019;14(10):1-18. doi:10.1371/journal.pone.0223796

23.	Zamboni S, Moschini M, Antonelli A, et al. How to improve patient selection for neoadjuvant chemotherapy in bladder cancer patients candidate for radical cystectomy and pelvic lymph node dissection. *World J Urol*. Published online August 2019. doi:10.1007/s00345-019-02916-2

24.	Nobles AL, Glenn JJ, Barnes LE. Identification of Inminent Suicide Risk Among Young Adults using Text Messages. *Proc SIGCHI Conf Hum Factor Comput Syst*. Published online 2019:1-22. doi:10.1145/3173574.3173987.Identification

25.	Tse R, Garland J, Kesha K, et al. Combining Postmortem Vitreous Sodium and Chloride and Lung-Body Ratio in Aiding the Diagnosing Saltwater Drowning. *Am J Forensic Med Pathol*. 2018;39(3):229-235. doi:10.1097/PAF.0000000000000410

26.	Sterling NW, Patzer RE, Mengyu D, et. a.Prediction of emergency department patient disposition based on natural language processing of triage notes. *Int J Med Inform*. 2019;129:184-188. doi:10.1016/j.ijmedinf.2019.06.008

27.	Perveen S, Shahbaz M, Keshavjee K, Guergachi A. A Systematic Machine Learning Based Approach for the Diagnosis of Non-Alcoholic Fatty Liver Disease Risk and Progression. *Sci Rep*. 2018;8(1):1-12. doi:10.1038/s41598-018-20166-x

28.	Pei Z, Liu J, Liu M, et al. Risk-Predicting Model for Incident of Essential Hypertension Based on Environmental and Genetic Factors with Support Vector Machine. *Interdiscip Sci Comput Life Sci*. 2018;10(1):126-130. doi:10.1007/s12539-017-0271-2

29.	Bouallègue F Ben, Mariano-Goulart D, Payoux P. Joint Assessment of Quantitative 18 F-Florbetapir and 18 F-FDG Regional Uptake Using Baseline Data from the ADNI. *J Alzheimer’s Dis*. 2018;62(1):399-408. doi:10.3233/JAD-170833

30.	Tsai TL, Huang MH, Lee CY, et. al. Data Science for Extubation Prediction and Value of Information in Surgical Intensive Care Unit. *J Clin Med*. 2019;8(10). doi:10.3390/jcm8101709

31.	Lopes RR, van Mourik MS, Schaft E V., et al. Value of machine learning in predicting TAVI outcomes. *Netherlands Hear J*. 2019;27(9):443-450. doi:10.1007/s12471-019-1285-7

32.	Maragatham G, Devi S. LSTM Model for Prediction of Heart Failure in Big Data. *J Med Syst*. 2019;43(5). doi:10.1007/s10916-019-1243-3

33.	Shao CY, Liu KC, Li CL, et al. C-reactive protein to albumin ratio is a key indicator in a predictive model for anastomosis leakage after esophagectomy: Application of classification and regression tree analysis. *Thorac Cancer*. 2019;10(4):728-737. doi:10.1111/1759-7714.12990

34.	Saccà V, Sarica A, Novellino F, et al. Evaluation of machine learning algorithms performance for the prediction of early multiple sclerosis from resting-state FMRI connectivity data. *Brain Imaging Behav*. 2019;13(4):1103-1114. doi:10.1007/s11682-018-9926-9

35.	Cearns M, Opel N, Clark S, et al. Predicting rehospitalization within 2 years of initial patient admission for a major depressive episode: a multimodal machine learning approach. *Transl Psychiatry*. 2019;9(1). doi:10.1038/s41398-019-0615-2

36.	Bouts MJRJ, van der Grond J, Vernooij MW, et al. Detection of mild cognitive impairment in a community-dwelling population using quantitative, multiparametric MRI-based classification. *Hum Brain Mapp*. 2019;40(9):2711-2722. doi:10.1002/hbm.24554

37.	Huben NB, Hussein AA, May PR, et al. Development of a Patient-Based Model for Estimating Operative Times for Robot-Assisted Radical Prostatectomy. *J Endourol*. 2018;32(8):730-736. doi:10.1089/end.2018.0249

38.	Hale AT, Stonko DP, Brown A, et al. Machine-learning analysis outperforms conventional statistical models and CT classification systems in predicting 6-month outcomes in pediatric patients sustaining traumatic brain injury. *Neurosurg Focus*. 2018;45(5):1-7. doi:10.3171/2018.8.FOCUS17773

39.	Salvatore C, Castiglioni I. A wrapped multi-label classifier for the automatic diagnosis and prognosis of Alzheimer’s disease. *J Neurosci Methods*. 2018;302:58-65. doi:10.1016/j.jneumeth.2017.12.016

40.	Yang H, Bath PA. The Use of Data Mining Methods for the Prediction of Dementia: Evidence from the English Longitudinal Study of Aging. *IEEE J Biomed Heal Informatics*. 2020;24(2):345-353. doi:10.1109/JBHI.2019.2921418

41.	Zhou M, Fukuoka Y, Goldberg K, Vittinghoff E, Aswani A. Applying machine learning to predict future adherence to physical activity programs. *BMC Med Inform Decis Mak*. 2019;19(1):0-11. doi:10.1186/s12911-019-0890-0

42.	Kang X, Liang Y, Wang S, et al. Prediction model comparison for gestational diabetes mellitus with macrosomia based on risk factor investigation. *J Matern Neonatal Med*. 2019;0(0):1-10. doi:10.1080/14767058.2019.1668922

43.	Sauer CM, Sasson D, Paik KE, et al. Feature selection and prediction of treatment failure in tuberculosis. *PLoS One*. 2018;13(11):1-14. doi:10.1371/journal.pone.0207491

44.	Thornblade LW, Flum DR, Flaxman AD. Predicting Future Elective Colon Resection for Diverticulitis Using Patterns of Health Care Utilization. *eGEMs (Generating Evid Methods to Improv patient outcomes)*. 2018;6(1):1. doi:10.5334/egems.193

45.	Lei VJ, Kennedy EH, Luong T, et al. Model performance metrics in assessing the value of adding intraoperative data for death prediction: Applications to noncardiac surgery. *Stud Health Technol Inform*. 2019;264:223-227. doi:10.3233/SHTI190216

46.	Lee SJ, Yu SH, Kim Y, et al. Development of integrated data and prediction system platform for the localized prostate cancer. *Stud Health Technol Inform*. 2019;264:1506-1507. doi:10.3233/SHTI190507

47.	Auffenberg GB, Ghani KR, Ramani S, et al. askMUSIC: Leveraging a Clinical Registry to Develop a New Machine Learning Model to Inform Patients of Prostate Cancer Treatments Chosen by Similar Men. *Eur Urol*. 2019;75(6):901-907. doi:10.1016/j.eururo.2018.09.050

48.	Wang Z, Wang B, Zhou Y, Li D, Yin Y. Weight-based multiple empirical kernel learning with neighbor discriminant constraint for heart failure mortality prediction. *J Biomed Inform*. 2020;101(November 2018):103340. doi:10.1016/j.jbi.2019.103340

49.	Nelson A, Herron D, Rees G, Nachev P. Predicting scheduled hospital attendance with artificial intelligence. *npj Digit Med*. 2019;2(1):1-7. doi:10.1038/s41746-019-0103-3

50.	Shibahara T, Ikuta S, Muragaki Y. Machine-Learning Approach for Modeling Myelosuppression Attributed to Nimustine Hydrochloride. *JCO Clin Cancer Informatics*. 2018;(2):1-21. doi:10.1200/cci.17.00022

51.	Liang S, Brown MRG, Deng W, et al. Convergence and divergence of neurocognitive patterns in schizophrenia and depression. *Schizophr Res*. 2018;192:327-334. doi:10.1016/j.schres.2017.06.004

52.	Chen R, Stewart WF, Sun J, Ng K, Yan X. Recurrent neural networks for early detection of heart failure from longitudinal electronic health record data: Implications for temporal modeling with respect to time before diagnosis, data density, data quantity, and data type. *Circ Cardiovasc Qual Outcomes*. 2019;12(10):1-15. doi:10.1161/CIRCOUTCOMES.118.005114

53.	Klang E, Barash Y, Soffer S, et al. Promoting head CT exams in the emergency department triage using a machine learning model. *Neuroradiology*. 2020;62(2):153-160. doi:10.1007/s00234-019-02293-y

54.	Fan Y, Li Y, Li Y, et al. Development and assessment of machine learning algorithms for predicting remission after transsphenoidal surgery among patients with acromegaly. *Endocrine*. 2020;67(2):412-422. doi:10.1007/s12020-019-02121-6

55.	Ferré Á, Poca MA, De La Calzada MD, et al. A conditional inference tree model for predicting sleep-related breathing disorders in patients with Chiari malformation type 1: Description and external validation. *J Clin Sleep Med*. 2019;15(1):89-99. doi:10.5664/jcsm.7578

56.	Zhang F, Zhang Y, Ke C, et al. Predicting ovarian cancer recurrence by plasma metabolic profiles before and after surgery. *Metabolomics*. 2018;14(5):1-9. doi:10.1007/s11306-018-1354-8

57.	Ferrer-Peña R, Calvo-Lobo C, Gómez M, Muñoz-García D. Prediction Model for Choosing Needle Length to Minimize Risk of Median Nerve Puncture With Dry Needling of the Pronator Teres. *J Manipulative Physiol Ther*. 2019;42(5):366-371. doi:10.1016/j.jmpt.2018.11.020

58.	Cameron JM, Butler HJ, Smith BR, et al. Developing infrared spectroscopic detection for stratifying brain tumour patients: Glioblastoma multiforme: Vs. lymphoma. *Analyst*. 2019;144(22):6736-6750. doi:10.1039/c9an01731c

59.	Harris AH, Kuo AC, Bowe T, Gupta S, Nordin D, Giori NJ. Prediction Models for 30-Day Mortality and Complications After Total Knee and Hip Arthroplasties for Veteran Health Administration Patients With Osteoarthritis. *J Arthroplasty*. 2018;33(5):1539-1545. doi:10.1016/j.arth.2017.12.003

60.	Kuo KM, Talley PC, Huang CH, Cheng LC. Predicting hospital-acquired pneumonia among schizophrenic patients: A machine learning approach. *BMC Med Inform Decis Mak*. 2019;19(1):1-8. doi:10.1186/s12911-019-0792-1

61.	Arai Y, Kondo T, Fuse K, et al. Using a machine learning algorithm to predict acute graft-versus-host disease following allogeneic transplantation. *Blood Adv*. 2019;3(22):3626-3634. doi:10.1182/bloodadvances.2019000934

62.	Paul JK, Iype T, R D, Hagiwara Y, Koh JEW, Acharya UR. Characterization of fibromyalgia using sleep EEG signals with nonlinear dynamical features. *Comput Biol Med*. 2019;111(April):103331. doi:10.1016/j.compbiomed.2019.103331

63.	Liu L, Yu Y, Fei Z, et al. An interpretable boosting model to predict side effects of analgesics for osteoarthritis. *BMC Syst Biol*. 2018;12(Suppl 6). doi:10.1186/s12918-018-0624-4

64.	Shappell C, Snyder A, Edelson DP, et. al. Predictors of In-hospital Mortality after rapid response team calls in a 274 hospital nationwide sample. *Crit Care Med*. 2018;176(10):139-148. doi:10.1097/CMM.00000000000002926

65.	Zhang R, Jing Y, Zhang H, et al. Comprehensive Evolutionary Analysis of the Major RNA-Induced Silencing Complex Members. *Sci Rep*. 2018;8(1):14189. doi:10.1038/s41598-018-32635-4

66.	Ge F, Li Y, Yuan M, Zhang J, Zhang W. Identifying predictors of probable posttraumatic stress disorder in children and adolescents with earthquake exposure: A longitudinal study using a machine learning approach. *J Affect Disord*. 2020;264(September 2019):483-493. doi:10.1016/j.jad.2019.11.079

67.	Dhondt B, De Bleser E, Claeys T, et al. Discovery and validation of a serum microRNA signature to characterize oligo- and polymetastatic prostate cancer: not ready for prime time. *World J Urol*. 2019;37(12):2557-2564. doi:10.1007/s00345-018-2609-8

68.	Rohaut B, Doyle KW, Reynolds AS, et al. Deep structural brain lesions associated with consciousness impairment early after hemorrhagic stroke. *Sci Rep*. 2019;9(1):1-9. doi:10.1038/s41598-019-41042-2

69.	Karnuta JM, Navarro SM, Haeberle HS, et al. Predicting Inpatient Payments Prior to Lower Extremity Arthroplasty Using Deep Learning: Which Model Architecture Is Best? *J Arthroplasty*. 2019;34(10):2235-2241.e1. doi:10.1016/j.arth.2019.05.048

70.	Wilson MB, Ali SA, Kovatch KJ, Smith JD, Hoff PT. Machine Learning Diagnosis of Peritonsillar Abscess. *Otolaryngol - Head Neck Surg (United States)*. 2019;161(5):796-799. doi:10.1177/0194599819868178

71.	Lu B, Fu L, Nie B, Peng Z, Liu H. A novel framework with high diagnostic sensitivity for lung cancer detection by electronic nose. *Sensors (Switzerland)*. 2019;19(23):1-29. doi:10.3390/s19235333

72.	Xu Y, Kong S, Cheung WY, et al. Development and validation of case-finding algorithms for recurrence of breast cancer using routinely collected administrative data. *BMC Cancer*. 2019;19(1):1-10. doi:10.1186/s12885-019-5432-8

73.	Cohen S, Fulcher BD, Rajaratnam SMW, et al. Sleep patterns predictive of daytime challenging behavior in individuals with low-functioning autism. *Autism Res*. 2018;11(2):391-403. doi:10.1002/aur.1899

74.	Wang Y, Liu W, Yu Y, et al. Prediction of the Depth of Tumor Invasion in Gastric Cancer: Potential Role of CT Radiomics. *Acad Radiol*. 2019;(6):1-8. doi:10.1016/j.acra.2019.10.020

75.	Mortezagholi A, Khosravizadehorcid O, Menhaj MB, Shafigh Y, Kalhor R. Make intelligent of gastric cancer diagnosis error in Qazvin’s medical centers: Using data mining method. *Asian Pacific J Cancer Prev*. 2019;20(9):2607-2610. doi:10.31557/APJCP.2019.20.9.2607

76.	Muehlematter UJ, Mannil M, Becker AS, et al. Vertebral body insufficiency fractures: detection of vertebrae at risk on standard CT images using texture analysis and machine learning. *Eur Radiol*. 2019;29(5):2207-2217. doi:10.1007/s00330-018-5846-8

77.	Faruqui SHA, Du Y, Meka R, et al. Development of a deep learning model for dynamic forecasting of blood glucose level for type 2 diabetes mellitus: Secondary analysis of a randomized controlled trial. *JMIR mHealth uHealth*. 2019;7(11):1-14. doi:10.2196/14452

78.	Molinari M, Ayloo S, Tsung A, et al. Prediction of Perioperative Mortality of Cadaveric Liver Transplant Recipients during Their Evaluations. *Transplantation*. 2019;103(10):E297-E307. doi:10.1097/TP.0000000000002810

79.	Jeon JP, Kim C, Oh BD, Kim SJ, Kim YS. Prediction of persistent hemodynamic depression after carotid angioplasty and stenting using artificial neural network model. *Clin Neurol Neurosurg*. 2018; 127-131. doi:10.1016/j.clineuro.2017.12.005

80.	Staartjes VE, Serra C, Muscas G, et al. Utility of deep neural networks in predicting gross-total resection after transsphenoidal surgery for pituitary adenoma: A pilot study. *Neurosurg Focus*. 2018;45(5):1-7. doi:10.3171/2018.8.FOCUS18243

81.	Lu CF, Hsu FT, Hsieh KLC, et al. Machine learning–based radiomics for molecular subtyping of gliomas. *Clin Cancer Res*. 2018;24(18):4429-4436. doi:10.1158/1078-0432.CCR-17-3445

82.	Park N, Kang E, Park M, et al. Predicting acute kidney injury in cancer patients using heterogeneous and irregular data. *PLoS One*. 2018;13(7):1-21. doi:10.1371/journal.pone.0199839

83.	Ballarini T, Mueller K, Albrecht F, et al. Regional gray matter changes and age predict individual treatment response in Parkinson’s disease. *NeuroImage Clin*. 2019;21(May 2018). doi:10.1016/j.nicl.2018.101636

84.	Chen D, Goyal G, Go RS, Parikh SA, Ngufor CG. Improved Interpretability of Machine Learning Model Using Unsupervised Clustering: Predicting Time to First Treatment in Chronic Lymphocytic Leukemia. *JCO Clin Cancer Informatics*. 2019;(3):1-11. doi:10.1200/cci.18.00137

85.	Malycha J, Farajidavar N, Pimentel MAF, et al. The effect of fractional inspired oxygen concentration on early warning score performance: A database analysis. *Resuscitation*. 2019;139(April 2019):192-199. doi:10.1016/j.resuscitation.2019.04.002

86.	Xie Z, Nikolayeva O, Luo J, Li D. Building risk prediction models for type 2 diabetes using machine learning techniques. *Prev Chronic Dis*. 2019;16(9):1-9. doi:10.5888/pcd16.190109

87.	Thanathornwong B. Bayesian-based decision support system for assessing the needs for orthodontic treatment. *Healthc Inform Res*. 2018;24(1):22-28. doi:10.4258/hir.2018.24.1.22

88.	Ngo CQ, Truong BCQ, Jones TW, Nguyen HT. Occipital EEG Activity for the Detection of Nocturnal Hypoglycemia. *Proc Annu Int Conf IEEE Eng Med Biol Soc EMBS*. 2018;2018-July:3862-3865. doi:10.1109/EMBC.2018.8513069

89.	Hyun SH, Ahn MS, Koh YW, et.al. A Machine-Learning Approach Using PET-Based Radiomics to Predict the Histological Subtypes of Lung Cancer. *Clin Nucl Med*. 2019;44(12):956-960. doi:10.1097/RLU.0000000000002810

90.	Rozet A, Kronish IM, Schwartz JE, Davidson KW. Using machine learning to derive just-in-time and personalized predictors of stress: Observational study bridging the gap between nomothetic and ideographic approaches. *J Med Internet Res*. 2019;21(4):1-16. doi:10.2196/12910

91.	Mellem MS, Liu Y, Gonzalez H, Kollada M, Martin WJ, Ahammad P. Machine Learning Models Identify Multimodal Measurements Highly Predictive of Transdiagnostic Symptom Severity for Mood, Anhedonia, and Anxiety. *Biol Psychiatry Cogn Neurosci Neuroimaging*. 2020;5(1):56-67. doi:10.1016/j.bpsc.2019.07.007

92.	Karhade A V., Ogink PT, Thio QCBS, et al. Development of machine learning algorithms for prediction of prolonged opioid prescription after surgery for lumbar disc herniation. *Spine J*. 2019;19(11):1764-1771. doi:10.1016/j.spinee.2019.06.002

93.	Zhang F, Tian S, Chen S, Ma Y, Li X, Guo X. Voxel-Based Morphometry: Improving the Diagnosis of Alzheimer’s Disease Based on an Extreme Learning Machine Method from the ADNI cohort. *Neuroscience*. 2019;414:273-279. doi:10.1016/j.neuroscience.2019.05.014

94.	Leightley D, Williamson V, Darby J, Fear NT. Identifying probable post-traumatic stress disorder: applying supervised machine learning to data from a UK military cohort. *J Ment Heal*. 2019;28(1):34-41. doi:10.1080/09638237.2018.1521946

95.	Mulder M, Nijland RH, van de Port IG, van Wegen EE, Kwakkel G. Prospectively Classifying Community Walkers After Stroke: Who Are They? *Arch Phys Med Rehabil*. 2019;100(11):2113-2118. doi:10.1016/j.apmr.2019.04.017

96.	Debédat J, Sokolovska N, Coupaye M, et al. Long-term Relapse of Type 2 Diabetes After Roux-en-Y Gastric Bypass: Prediction and clinical relevance. *Diabetes Care*. 2018;41(10):2086-2095. doi:10.2337/dc18-0567

97.	Alcantud JCR, Varela G, Santos-Buitrago B, Santos-García G, Jiménez MF. Analysis of survival for lung cancer resections cases with fuzzy and soft set theory in surgical decision making. *PLoS One*. 2019;14(6):1-17. doi:10.1371/journal.pone.0218283

98.	Sandström A, Snowden JM, Höijer J, Bottai M, Wikström AK. Clinical risk assessment in early pregnancy for preeclampsia in nulliparous women: A population based cohort study. *PLoS One*. 2019;14(11):1-16. doi:10.1371/journal.pone.0225716

99.	Xiao C, Liu Y, Feng DD, Wang X. Key Marker Selection for the Detection of Early Parkinson’ s Disease using Importance-Driven Models. *Conf Proc . Annu Int Conf IEEE Eng Med Biol Soc IEEE Eng Med Biol Soc Annu Conf*. 2018;2018:6100-6103. doi:10.1109/EMBC.2018.8513564

100.	Cooper JN, Minneci PC, Deans KJ. Postoperative neonatal mortality prediction using superlearning. *J Surg Res*. 2018;221:311-319. doi:10.1016/j.jss.2017.09.002

101.	Rau CS, Kuo PJ, Chien PC, Huang CY, Hsieh HY, Hsieh CH. Mortality prediction in patients with isolated moderate and severe traumatic brain injury using machine learning models. *PLoS One*. 2018;13(11):1-12. doi:10.1371/journal.pone.0207192

102.	Anand RS, Stey P, Jain S, et al. Predicting Mortality in Diabetic ICU Patients Using Machine Learning and Severity Indices. *AMIA Jt Summits Transl Sci proceedings AMIA Jt Summits Transl Sci*. 2018;2017:310-319.

103.	Aperstein Y, Cohen L, Bendavid I, et al. Improved ICU mortality prediction based on SOFA scores and gastrointestinal parameters. *PLoS One*. 2019;14(9):1-11. doi:10.1371/journal.pone.0222599

104.	Park J, Kim JW, Ryu B, Heo E, Jung SY, Yoo S. Patient-level prediction of cardio-cerebrovascular events in hypertension using nationwide claims data. *J Med Internet Res*. 2019;21(2). doi:10.2196/11757

105.	Gökçay D, Eken A, Baltaci S. Binary Classification Using Neural and Clinical Features: An Application in Fibromyalgia with Likelihood-Based Decision Level Fusion. *IEEE J Biomed Heal Informatics*. 2019;23(4):1490-1498. doi:10.1109/JBHI.2018.2844300

106.	Gupta R, Gupta S, Chauhan L. Predicting visual outcome after open globe injury using classification and regression tree model: the Moradabad ocular trauma study. *Can J Ophthalmol*. 2019;54(4):473-478. doi:10.1016/j.jcjo.2018.08.004

107.	Butt AH, Rovini E, Dolciotti C, et al. Objective and automatic classifcation of Parkinson disease with Leap Motion controller. *Biomed Eng Online*. 2018;17(1):1-21. doi:10.1186/s12938-018-0600-7

108.	Hunter-Zinck HS, Peck JS, Strout TD, Gaehde SA. Predicting emergency department orders with multilabel machine learning techniques and simulating effects on length of stay. *J Am Med Informatics Assoc*. 2019;26(12):1427-1436. doi:10.1093/jamia/ocz171

109.	Kilic A, Goyal A, Miller JK, et al. Predictive Utility of a Machine Learning Algorithm in Estimating Mortality Risk in Cardiac Surgery. *Ann Thorac Surg*. Published online 2020. doi:10.1016/j.athoracsur.2019.09.049

110.	Campillo-Artero C, Serra-Burriel M, Calvo-Pérez A. Predictive modeling of emergency cesarean delivery. *PLoS One*. 2018;13(1):1-14. doi:10.1371/journal.pone.0191248

111.	Zhang ML, Guo AX, Kadauke S, Dighe AS, Baron JM, Sohani AR. Machine Learning Models Improve the Diagnostic Yield of Peripheral Blood Flow Cytometry. *Am J Clin Pathol*. 2020;153(2):235-242. doi:10.1093/ajcp/aqz150

112.	Hong WS, Haimovich AD, Taylor RA. Predicting hospital admission at emergency department triage using machine learning. *PLoS One*. 2018;13(7):1-13. doi:10.1371/journal.pone.0201016

113.	Beauchet O, Noublanche F, Simon R, et. al. Falls risk prediction for older inpatients in acute care medical wards: is there an interest to combine an early nurse assessment and the artificial neural network analysis? *J Nutr Health Aging*. 2018. doi:10.1007/s12603-017-0950-z

114.	Ma Z, Wang P, Gao Z, Wang R, Khalighi K. Ensemble of machine learning algorithms using the stacked generalization approach to estimate the warfarin dose. *PLoS One*. 2018;13(10):1-12. doi:10.1371/journal.pone.0205872

115.	Veeranki SPK, Hayn D, Jauk S, et al. An improvised classification model for predicting delirium. *Stud Health Technol Inform*. 2019;264:1566-1567. doi:10.3233/SHTI190537

116.	Maharlou H, Niakan Kalhori SR, Shahbazi S, Ravangard R. Predicting length of stay in intensive care units after cardiac surgery: Comparison of artificial neural networks and adaptive neuro-fuzzy system. *Healthc Inform Res*. 2018;24(2):109-117. doi:10.4258/hir.2018.24.2.109

117.	Castillo-Olea C, B G-ZS, C CL, Zuniga C. Automatic Classification of Sarcopenia Level in Older Adults: A Case Study at Tijuana General Hospital. *Int J Environ Res Public Health*. 2019;16(18). doi:10.3390/ijerph16183275

118.	Sa-ngamuang C, Haddawy P, Luvira V, et al. Accuracy of dengue clinical diagnosis with and without NS1 antigen rapid test: Comparison between human and Bayesian network model decision. *PLoS Negl Trop Dis*. 2018;12(6):1-14. doi:10.1371/journal.pntd.0006573

119.	Talaei-Khoei A, Wilson JM. Identifying people at risk of developing type 2 diabetes: A comparison of predictive analytics techniques and predictor variables. *Int J Med Inform*. 2018;119(January):22-38. doi:10.1016/j.ijmedinf.2018.08.008

120.	Bertsimas D, Dunn J, Velmahos GC, Kaafarani HMA. Surgical Risk Is Not Linear: Derivation and Validation of a Novel, User-friendly, and Machine-learning-based Predictive OpTimal Trees in Emergency Surgery Risk (POTTER) Calculator. *Ann Surg*. 2018;268(4):574-583. doi:10.1097/SLA.0000000000002956

121.	Liu C, Qi L, Feng QX, Sun SW, Zhang YD, Liu XS. Performance of a machine learning-based decision model to help clinicians decide the extent of lymphadenectomy (D1 vs. D2) in gastric cancer before surgical resection. *Abdom Radiol*. 2019;44(9):3019-3029. doi:10.1007/s00261-019-02098-w

122.	Luo G, Stone BL, Nkoy FL, He S, Johnson MD. Predicting appropriate hospital admission of emergency department patients with bronchiolitis: Secondary analysis. *JMIR Med Informatics*. 2019;7(1):1-15. doi:10.2196/12591

123.	Meena K, Tayal DK, Gupta V, Fatima A. Using classification techniques for statistical analysis of Anemia. *Artif Intell Med*. 2019;94(August 2018):138-152. doi:10.1016/j.artmed.2019.02.005

124.	Takeuchi M, Kawakubo H, Mayanagi S, et al. Postoperative Pneumonia is Associated with Long-Term Oncologic Outcomes of Definitive Chemoradiotherapy Followed by Salvage Esophagectomy for Esophageal Cancer. *J Gastrointest Surg*. 2018;22(11):1881-1889. doi:10.1007/s11605-018-3857-z

125.	Kiiski H, Jollans L, SO D, et al. Machine Learning EEG to Predict Cognitive Functioning and Processing Speed Over a 2-Year Period in Multiple Sclerosis Patients and Controls. *Brain Topogr*. 2018;31(3):346-363. doi:10.1007/s10548-018-0620-4

126.	Hasnain Z, Mason J, Gill K, et al. Machine learning models for predicting post-cystectomy recurrence and survival in bladder cancer patients. *PLoS One*. 2019;14(2):1-15. doi:10.1371/journal.pone.0210976

127.	Dean J, Wong K, Gay H, et al. Incorporating spatial dose metrics in machine learning-based normal tissue complication probability (NTCP) models of severe acute dysphagia resulting from head and neck radiotherapy. *Clin Transl Radiat Oncol*. 2018;8:27-39. doi:10.1016/j.ctro.2017.11.009

128.	Cheung M, Cobb AN, Kuo PC. Predicting burn patient mortality with electronic medical records. *Surg (United States)*. 2018;164(4):839-847. doi:10.1016/j.surg.2018.07.010

129.	Gowin JL, Ernst M, Ball T, et al. Using neuroimaging to predict relapse in stimulant dependence: A comparison of linear and machine learning models. *NeuroImage Clin*. 2019;21(January):101676. doi:10.1016/j.nicl.2019.101676

130.	Rojas JC, Carey KA, Edelson DP, Venable LR, Howell MD, Churpek MM. Predicting intensive care unit readmission with machine learning using electronic health record data. *Ann Am Thorac Soc*. 2018;15(7):846-853. doi:10.1513/AnnalsATS.201710-787OC

131.	Wei R, Lin K, Yan W, et al. Computer-Aided Diagnosis of Pancreas Serous Cystic Neoplasms: A Radiomics Method on Preoperative MDCT Images. *Technol Cancer Res Treat*. 2019;18:1533033818824339. doi:10.1177/1533033818824339

132.	Balani J, Hyer SL, Shehata H, Mohareb F. Visceral fat mass as a novel risk factor for predicting gestational diabetes in obese pregnant women. *Obstet Med*. 2018;11(3):121-125. doi:10.1177/1753495X17754149

133.	Gao L, Smielewski P, Li P, Czosnyka M, Ercole A. Signal Information Prediction of Mortality Identifies Unique Patient Subsets after Severe Traumatic Brain Injury: A Decision-Tree Analysis Approach. *J Neurotrauma*. doi:10.1089/neu.2019.6631

134.	Garcia-Arce A, Rico F, Zayas-Castro JL. Comparison of Machine Learning Algorithms for the Prediction of Preventable Hospital Readmissions. *J Healthc Qual*. 2018;40(3):129-138. doi:10.1097/JHQ.0000000000000080

135.	Papini S, Pisner D, Shumake J, et al. Ensemble machine learning prediction of posttraumatic stress disorder screening status after emergency room hospitalization. *J Anxiety Disord*. 2018;60(October):35-42. doi:10.1016/j.janxdis.2018.10.004

136.	Jang BS, Jeon SH, Kim IH, Kim IA. Prediction of Pseudoprogression versus Progression using Machine Learning Algorithm in Glioblastoma. *Sci Rep*. 2018;8(1):1-9. doi:10.1038/s41598-018-31007-2

137.	Cosgriff C V., Celi LA, Ko S, et al. Developing well-calibrated illness severity scores for decision support in the critically ill. *npj Digit Med*. 2019;2(1). doi:10.1038/s41746-019-0153-6

138.	Kim H, Lee SH, Lee SE, Hong S, Kang HJ, Kim N. Depression prediction by using ecological momentary assessment, actiwatch data, and machine learning: Observational study on older adults living alone. *JMIR mHealth uHealth*. 2019;7(10). doi:10.2196/14149

139.	Lötsch J, Sipilä R, Tasmuth T, et al. Machine-learning-derived classifier predicts absence of persistent pain after breast cancer surgery with high accuracy. *Breast Cancer Res Treat*. 2018;171(2):399-411. doi:10.1007/s10549-018-4841-8

140.	Spyroglou II, Spöck G, Rigas AG, Paraskakis EN. Evaluation of Bayesian classifiers in asthma exacerbation prediction after medication discontinuation. *BMC Res Notes*. 2018;11(1):18-23. doi:10.1186/s13104-018-3621-1

141.	Goudey B, Fung BJ, Schieber C, et al. A blood-based signature of cerebrospinal fluid Aβ 1–42 status. *Sci Rep*. 2019;9(1):0-12. doi:10.1038/s41598-018-37149-7

142.	Facciorusso A, Del Prete V, Antonino M, Buccino VR, Muscatiello N. Response to repeat echoendoscopic celiac plexus neurolysis in pancreatic cancer patients: A machine learning approach. *Pancreatology*. 2019;19(6):866-872. doi:10.1016/j.pan.2019.07.038

143.	Kim DW, Kim H, Nam W, Kim HJ, Cha IH. Machine learning to predict the occurrence of bisphosphonate-related osteonecrosis of the jaw associated with dental extraction: A preliminary report. *Bone*. 2018;116(January):207-214. doi:10.1016/j.bone.2018.04.020

144.	Karhade A V., Shah AA, Bono CM, et al. Development of machine learning algorithms for prediction of mortality in spinal epidural abscess. *Spine J*. 2019;19(12):1950-1959. doi:10.1016/j.spinee.2019.06.024

145.	Tu W, Chen PA, Koenig N, et al. Machine learning models reveal neurocognitive impairment type and prevalence are associated with distinct variables in HIV/AIDS. *J Neurovirol*. 2020;26(1):41-51. doi:10.1007/s13365-019-00791-6

146.	Van Steenkiste T, Ruyssinck J, De Baets L, et al. Accurate prediction of blood culture outcome in the intensive care unit using long short-term memory neural networks. *Artif Intell Med*. 2019;97(April 2017):38-43. doi:10.1016/j.artmed.2018.10.008

147.	Paliwal N, Jaiswal P, Tutino VM, et al. Outcome prediction of intracranial aneurysm treatment by flow diverters using machine learning. *Neurosurg Focus*. 2018;45(5):1-10. doi:10.3171/2018.8.FOCUS18332

148.	Bronsert M, Singh AB, Henderson WG, Hammermeister K, Meguid RA, Colborn KL. Identification of postoperative complications using electronic health record data and machine learning. *Am J Surg*. 2020;220(1):114-119. doi:10.1016/j.amjsurg.2019.10.009

149.	Eill A, Jahedi A, Gao Y, et al. Functional Connectivities Are More Informative Than Anatomical Variables in Diagnostic Classification of Autism. *Brain Connect*. 2019;9(8):604-612. doi:10.1089/brain.2019.0689

150.	Cook F, Lobo D, Martin M, et al. Prospective validation of a new airway management algorithm and predictive features of intubation difficulty. *Br J Anaesth*. 2019;122(2):245-254. doi:10.1016/j.bja.2018.09.021

151.	Eggleston B, Dismuke-Greer CE, Pogoda TK, et al. A prediction model of military combat and training exposures on VA service-connected disability: a CENC study. *Brain Inj*. 2019;33(13-14):1602-1614. doi:10.1080/02699052.2019.1655793

152.	Villarreal YR, Suchting R, Klawans MR, et al. Predicting HCV Incidence in Latinos with High-Risk Substance Use: A Data Science Approach. *Soc Work Public Health*. 2019;34(7):606-615. doi:10.1080/19371918.2019.1635948
